# Reconstructing the early spatial spread of pandemic respiratory viruses in the United States

**DOI:** 10.1101/2025.11.24.25340792

**Authors:** Renquan Zhang, Rui Deng, Sitong Liu, Qing Yao, Jeffrey Shaman, Bryan T Grenfell, Cécile Viboud, Sen Pei

## Abstract

Understanding the geographic spread of emerging respiratory viruses is critical for pandemic preparedness, yet the early spatiotemporal dynamics of the 2009 H1N1 pandemic influenza and SARS-CoV-2 in the United States (US) remain unclear. While mobility and genomic data have revealed important aspects of pandemic spatial spread, several key questions remain: Did the two pandemics follow similar spatial transmission routes? How rapidly did they spread across the US? What role did stochastic processes play in early spatial transmission? To address these questions, we integrated high-resolution disease data with a robust, data-efficient inference framework combining air travel, commuting flows, and pathogen superspreading potentials to reconstruct their spatial spread across US metropolitan areas. The two pandemics exhibited distinct transmission pathways across locations; however, both pandemics established local circulation in most metropolitan areas within weeks, driven by several shared transmission hubs. Early spatial spread was more strongly associated with air travel than with commuting, though stochastic dynamics introduced substantial uncertainty in transmission routes, creating challenges for timely detection and control. Simulations indicate that broad wastewater surveillance coverage beyond top transmission hubs coupled with effective infection control may slow initial spatial expansion. Our findings highlight the rapid, stochastic spread of pandemic respiratory pathogens and the difficulties of early outbreak containment.

## Introduction

Emerging respiratory viruses pose a persistent threat to human health and society. Over the past two decades, two global respiratory pandemics have occurred: A/H1N1 influenza (H1N1pdm influenza) (1) and COVID-19 (2). Other novel zoonotic pathogens, such as severe acute respiratory syndrome coronavirus (SARS-CoV) (3) and Middle East respiratory syndrome coronavirus (MERS-CoV) (4), have caused severe epidemics, and the recent circulation of A/H5N1 avian influenza has further heightened concerns that another pandemic is imminent (5).

Understanding the geographic spread of novel respiratory pathogens is essential for effective pandemic preparedness and response. Extensive research has demonstrated the critical role of human mobility in shaping the spatial spread of respiratory diseases (6–15). Airline travel drives the global, long-range spread of infectious diseases (9–11), and short-range commuting further disseminates viruses to suburban and rural populations (6, 7). More recent studies have combined genomic and mobility data to elucidate the transmission of SARS-CoV-2 at the global, regional, and local scales (16–27). In the United States (US), research has examined factors influencing the spatial spread of influenza (28–30) and understand the early transmission of SARS-CoV-2 using data-driven models (31–34) and genomic data from specific regions (20–23). However, due to severe underreporting and a lack of high-resolution disease data, the early spatiotemporal dynamics of SARS-CoV-2 and H1N1pdm influenza in the US at the sub-state level remains poorly understood.

Here, leveraging city-level influenza-like illness (ILI) records from medical claims (35) and estimated county-level COVID-19 infections accounting for underreporting (31) (Materials and Methods, Fig. S1), we developed an inference framework to reconstruct the transmission pathways of H1N1pdm influenza and SARS-CoV-2 across US Metropolitan Statistical Areas (MSAs). Each MSA is defined as a densely populated urban center with close social and economic ties throughout the region, measured by commuting and employment (36). Due to frequent population mixing, MSAs serve as natural geographical units for characterizing the spatial spread of respiratory pathogens. The reconstructed transmission networks among MSAs allow comparison of the spatial spread patterns of the two viruses and reveal the rapid geographical expansion of the last two pandemics.

## Results

### Quantifying the uncertainty of early spatial spread

During the early stage of a respiratory viral pandemic, the spatial spread of a pathogen proceeds stochastically due to factors such as the randomness of human mobility trajectories and individual variation in infectiousness (e.g., superspreading events (37)). As a consequence, the establishment of sustained local transmission may require multiple introductions of infectious individuals (37). Although mobility flow drives the overall pattern of infection introductions, the realized transmission network is only one possible outcome of this stochastic process.

To quantify the uncertainty of these early spatial dynamics, we propose a process-based stochastic transmission model that incorporated inter-MSA air travel, commuting flows, and pathogen superspreading potential (Materials and Methods, Supplementary Information, Fig. S2). We simulated the outbreak of a hypothetical novel respiratory virus originating from Minnesota (Supplementary Information). For each MSA, we estimated the onset time of local transmission (38) (Fig. S3) and identified its infection source based on the number of infections introduced from other MSAs (Materials and Methods). A transmission link was defined from an identified infection source to a focal MSA and was categorized based on the mobility mode (commuting or flight) that contributed the most introductions. The simulated transmission network exhibited a hub-and-spoke structure (Fig. 1A, Figs. S4-S5), in agreement with previous simulation studies (6, 7).

**Figure 1.**
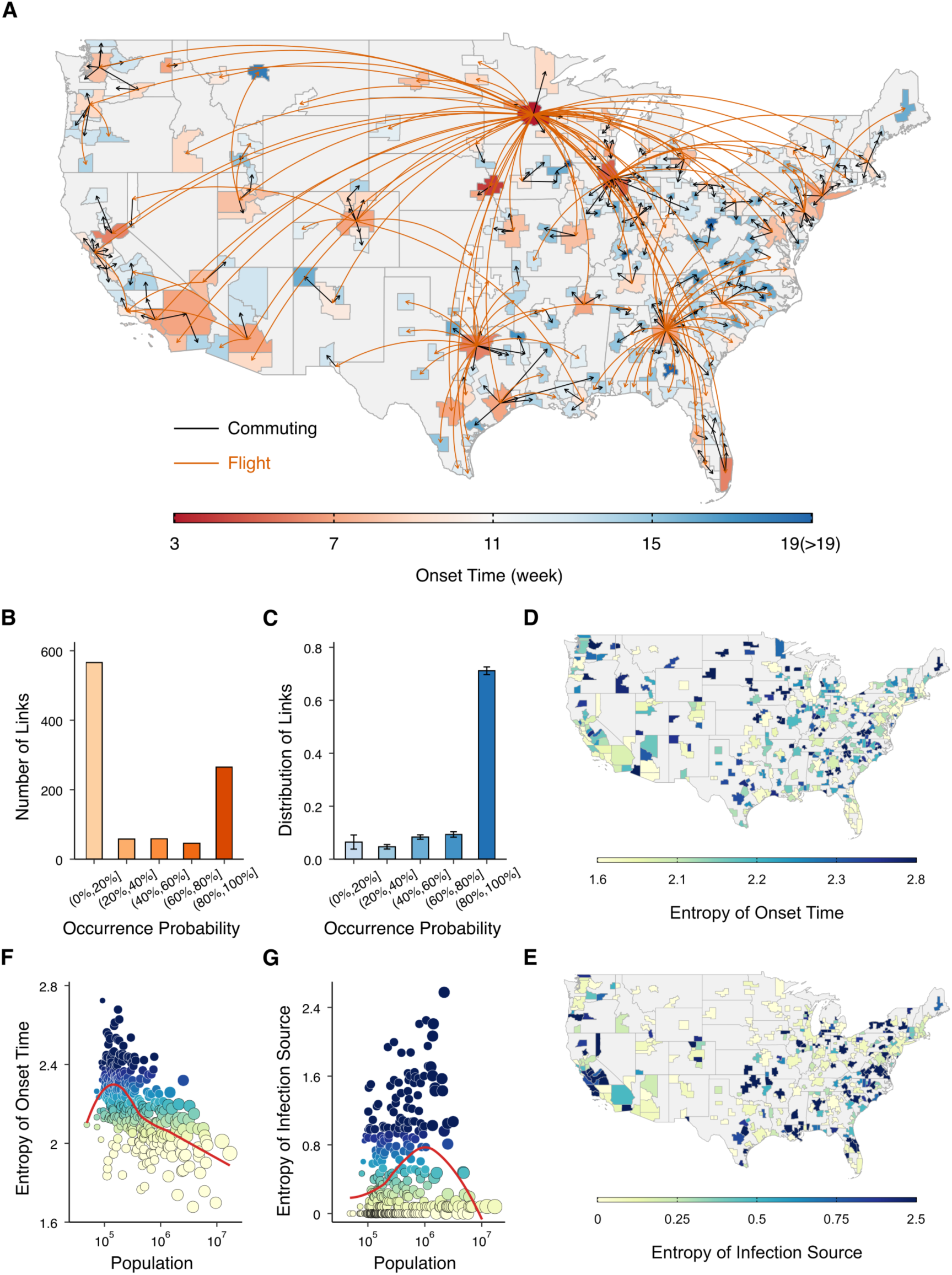
Simulating the early spatial spread of novel respiratory viruses. (**A**). The transmission network across the US MSAs in a single simulation. Onset times of local transmission in MSAs are shown by the color. Arrows represent transmission via commuting or flights, distinguished by color. (**B**). Numbers of transmission links that occurred with different probabilities in 100 independent simulations. Transmission links were classified into five categories based on their occurrence probabilities. (**C**). The distribution of transmission links generated within a single realization over the five categories. Error bars show the standard deviations obtained from 100 simulations. The entropy of local onset time (**D**) and infection source (**E**) for MSAs in 100 independent simulations are shown. Relationships between the population of MSAs and entropy are presented for local onset time (**F**) and infection source (**G**), respectively. Node size represents population size and color shows entropy. Red lines are curves fitting to the data using locally estimated scatterplot smoothing.

To explore the variability of transmission network structure, we performed 100 independent simulations and grouped transmission links that occurred at least once into five categories based on their occurrence probabilities. The beginning of an outbreak was defined as the onset time at the epidemic origin. Among the 100 independent realizations, the beginning of the outbreak was observed after 3 (95% CI [2, 5]) weeks of the simulation, when 670 (95% CI [406, 967]) infections had occurred at the epidemic origin. Among a total of 994 distinct transmission links, 566 (56.9%) links appeared less than 20% of the time, suggesting considerable variation in the transmission networks (Fig. 1B). Within a single realization, 71.1% (95% CI [68.5% - 73.5%]) of transmission links were stable, occurring in over 80% of simulations (Fig. 1C). Further simulations showed that the variability of the transmission network structure increases with lower virus transmissibility and higher superspreading potentials (Fig. S6).

We further quantified the stochasticity of the early spatiotemporal dynamics using the entropy of onset time and infection source: 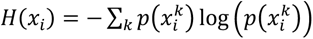, where 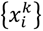 is the set of distinct onset dates or infection sources for location 𝑖, 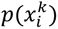 is the probability of 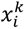 in the simulations, and the base of the logarithm is 𝑒 (Supplementary Information). A higher entropy indicates a less predictable quantity with a larger uncertainty. The entropy of onset time and infection source varied over a wide range across MSAs (Fig. 1D, E). The local onset time and infection source in many MSAs were intrinsically hard to predict, even when the same model and initial conditions were used. In general, the onset time for MSAs with a smaller population was less predictable (Fig. 1F). The uncertainty of the infection source was highest for MSAs with an intermediate population size (Fig. 1G), where infections could be introduced via multiple potential sources.

### An ensemble inference framework to reconstruct early spatial spread

To infer transmission across MSAs, it is essential to understand if infections in one location affect the epidemic in another place. Many approaches exist to detect such directional dynamical coupling between time series, such as Granger causality (39), transfer entropy (40), and convergent cross mapping (41). However, these techniques were not specifically designed for epidemic time series and cannot explicitly incorporate mechanistic understanding of spatial spread dynamics. Moreover, to ensure high inference performance, these approaches may require a large amount of training data (42), which are typically not available for disease surveillance, particularly early in an outbreak. To overcome these challenges, we developed a prediction-based inference framework tailored to the spatial spread of infectious diseases, incorporating prior knowledge of the connectivity between locations and nonlinear, stochastic transmission dynamics (Materials and Methods, Supplementary Information, Fig. S7).

We validated the inference framework using the simulated outbreak in Fig. 1A, for which we know both the realized, ground-truth transmission network and the model that generated the outbreak. Due to stochasticity in the transmission model, independent realizations of the inference procedure may result in different reconstructed transmission networks. To capture this uncertainty, 100 realizations of the inference were performed. Individually, the inference algorithm achieved a precision of 79.3% (95% CI [75.8% - 82.4%]) and a recall of 78.2% ([74.7% - 81.3%]) in identifying ground-truth transmission links (Fig. 2A). While over 600 transmission links only appeared in less than 20% of inference results, 240 transmission links were identified in over 80% of the inferred transmission networks (Fig. 2B). More importantly, the inference accuracy for these transmission links increased substantially with their occurrence probabilities among independent realizations (Fig. 2C). We examined inference performance using different thresholds of the occurrence probability to define transmission links. For this specific example, a threshold of 35% achieved the best overall performance, balancing a precision of 86.4% with a recall of 88.6% (Fig. 2D-E). The ensemble inference outperformed each individual inference in Fig. 2A, highlighting the benefit of ensembling outcomes from multiple realizations. The inference performance was robust to the selection of epidemic origins, misspecification of the underlying transmission model, and missingness of disease data in many locations (Supplementary Information, Figs. S8-S9).

**Figure 2.**
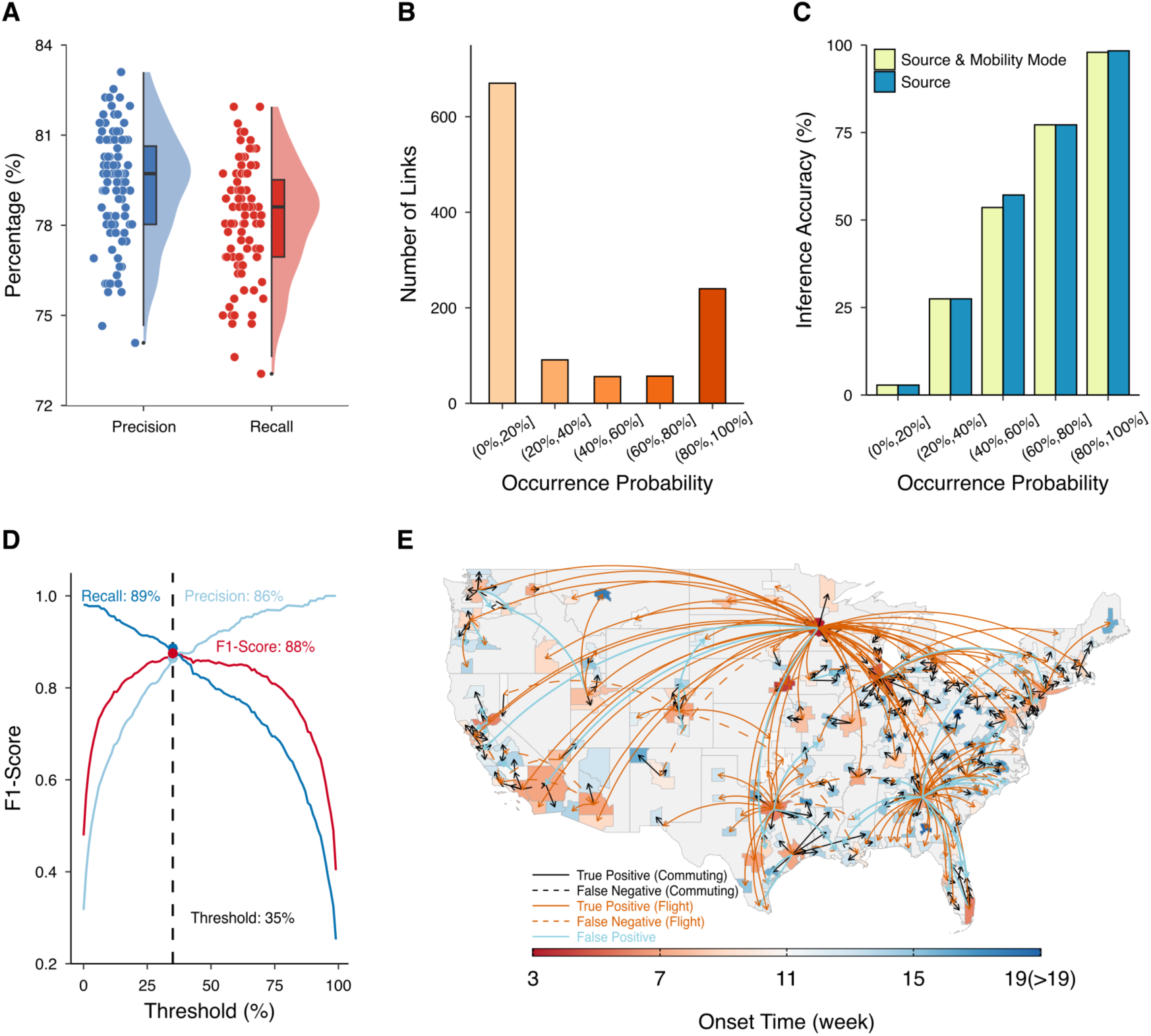
Inferring the spatial spread of a simulated outbreak. (**A**). Precision and recall for the results from 100 independent realizations of inference. Each dot represents the performance for one realization. Boxes show the medians and interquartile of the distributions and whiskers show 95% CIs. An inferred transmission link is defined as accurate if both the infection source and mobility mode are correct. (**B**). Numbers of transmission links that appeared with different probabilities in 100 realizations of inference. (**C**). The accuracy of inferred transmission links with different occurrence probabilities in the inference. Blue bars show the accuracy for identifying infection sources and yellow bars show the accuracy for additionally finding the correct mobility mode. (**D**). The performance of the ensemble inference using different thresholds of the occurrence probability to define transmission links. The F1-score is the harmonic mean of precision and recall, a metric to evaluate the overall performance of the inference. A threshold of 35% achieved the highest F1-score, with a precision of 86% and a recall of 89%. (**E**). The inferred transmission network using a threshold of 35% in the ensemble inference. True positives, false negatives, and false positives are shown using different colors and line types. Results are shown for both commuting and flight. The color of MSAs represents the onset time of local transmission.

To assess whether the inference algorithm can identify multiple introductions, we performed inference on an outbreak starting simultaneously from five outbreak origins – 1) Mobile, AL MSA; 2) Asheville, NC MSA; 3) San Francisco-Oakland-Fremont, CA MSA; 4) Minneapolis-St. Paul- Bloomington, MN-WI MSA; 5) New York-Northern New Jersey-Long Island, NY-NJ-PA MSA (Fig. S10). These locations include both metropolitan areas with strong inter-MSA mobility flows (San Francisco, Minneapolis, New York) and areas that are relatively isolated from other MSAs (Mobile, Asheville). In the inference, we selected the location with the earliest onset time as the sole outbreak origin and let the inference detect other origins. Among 100 independent inferences, Minneapolis was always selected as an origin as it had the earliest onset time. Other origins with less inter-MSA mobility were more likely to be captured. For instance, Mobile and Asheville were identified as outbreak origins in 95 and 94 reconstructed transmission networks, respectively. In contrast, outbreaks in San Francisco and New York were always inferred to be introduced from Minneapolis. Indeed, well-connected outbreak origins are inherently difficult to identify using mobility data alone, because their outbreaks may be explained by introductions from other MSAs with early onset. Additional data sources, such as genome sequences, are thus needed to further support outbreak origin inference.

To establish a “null model” for inference performance on any network structure, we performed inference on two randomized mobility networks. In the first, we rewired each commuting or flight link to a randomly selected destination, keeping the number of travelers for each link; in the second, we additionally assigned the same number of travelers to commuting or flight links from each MSA, removing the heterogeneity in mobility intensity for each location. We simulated an outbreak originating from Minnesota and performed inference using the randomized mobility networks. For the first randomization, the precision and recall of individual inference declined to 66.4% (95% CI [63.3%, 69.6%]) and 64.7% ([61.6%, 67.7%]) (Fig. S11). The ensemble inference had a precision of 73.0% with a recall of 76.7%. For the second randomization, the precision and recall of individual inference further dropped to 62.3% (95% CI [59.1%, 65.6%]) and 61.3% ([58.1%, 64.6%]), leading to a precision of 74.3% with a recall of 66.6% for the ensemble inference (Fig. S12). This comparison indicates that the real-world mobility network structure and mobility heterogeneity contain useful information that enables more accurate inference of transmission networks.

### The spatial transmission of SARS-CoV-2

We first applied the inference approach to reconstruct the transmission network of SARS-CoV-2 in the US from February 21^st^ to May 10^th^, 2020. Due to limited testing capacity in the early stage of the pandemic and the existence of asymptomatic and mild infections, COVID-19 case data were severely underreported. To address this issue, we used the daily new infections (both reported and unreported cases) estimated using a dynamic data-driven transmission model that accounted for time-varying ascertainment rates and reporting delays (31). The estimated infection numbers were validated independently by serological data collected at different locations and time points (31). This model was built at the US county level with inter-county mobility informed by the same commuting flows adjusted by real-time mobile phone-derived foot-traffic data. The estimated county-level infection numbers were aggregated to the MSA level to support inference. Informed by the estimated onset times and the genomic evidence for the introduction of SARS-CoV-2 into the US (20), Seattle and New York were selected as the infection origins in the inference. We performed 100 independent inferences using an optimal threshold tailored to the characteristics of SARS-CoV-2 (Materials and Methods, Supplementary Information, Fig. S9).

The inferred transmission network of SARS-CoV-2 consisted of 304 links with a hub-and-spoke structure (Fig. 3A-B, Fig. S13). Seattle and New York drove the long-range, national spread of the virus, mainly through air travel. Regional hubs such as Chicago, Atlanta, New Orleans, and San Francisco further propelled the dissemination to nearby MSAs. Most inter-MSA transmission links occurred during the two weeks from February 28^th^ to March 12^th^, 2020 (Fig. 3B), before the announcement of a nationwide emergency on March 13^th^, 2020 (43). Transmission networks inferred using other thresholds (Fig. S14), different infection origins (Fig. S15), and a shorter immunity duration (Fig. S16) remained structurally similar.

**Figure 3.**
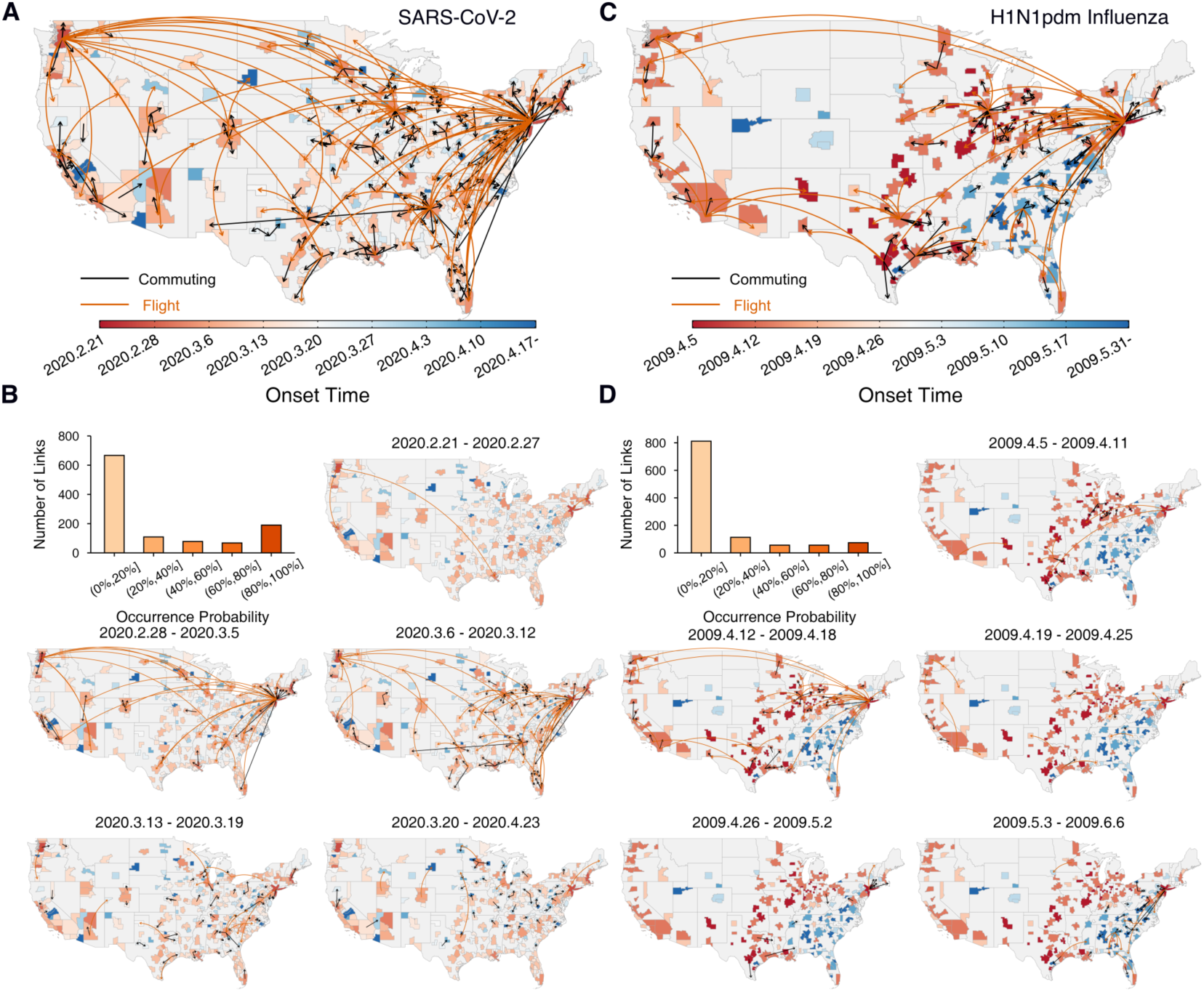
Inference of the early spatial spread of SARS-CoV-2 and H1N1pdm influenza. (**A**). The inferred transmission network for SARS-CoV-2. We performed 100 independent realizations of inference and selected 304 transmission links that appeared in over 46% of inference results. (B). The inferred transmission links of SARS-CoV-2 occurred in different weeks after February 21^st^, 2020. The bar plot shows the numbers of transmission links with different occurrence probabilities in the inference results. (**C**). The inferred transmission network for H1N1pdm influenza. 168 transmission links were selected using a threshold of 42% for the occurrence probability in 100 inference realizations. (**D**). The inferred transmission links of H1N1pdm influenza occurred in different weeks after April 5^th^, 2009. The bar plot represents the numbers of transmission links with different occurrence probabilities in the inference.

The inference results agreed with several findings from independent genomic studies performed in specific regions of the US. Phylogenetic analysis of SARS-CoV-2 genomes from Northern California suggested that the virus was likely disseminated from Washington state to California (22). Our inference identified a transmission link from the Seattle area to San Franscisco in February 2020 (Fig. 3B). Another study found that SARS-CoV-2 circulating in Mississippi and Alabama could be traced back to New Orleans (21); in agreement with this finding, we inferred transmission links from New Orleans to nearby MSAs through commuting. Additionally, phylogenetic analysis found that SARS-CoV-2 was introduced into Louisiana via domestic travel and occurred prior to the Mardi Gras festival on February 25^th^, 2020 (21). Accordingly, we inferred a transmission link from Washington state to New Orleans in the week of February 21^st^ via air travel (Fig. 3B), suggesting introductions prior to February 21^st^. Different from our inference, the genomic data suggested that SARS-CoV-2 in Louisiana may have originated from Texas; however, the same study also estimated a high importation risk from Washington state to New Orleans based on the estimated number of infectious travelers (21).

### The spatial transmission of H1N1pdm influenza

We next reconstructed the transmission network of H1N1pdm influenza during the 2009 spring wave. Although the spatial spread during its autumn wave has been studied extensively (28, 30, 38), the spread of the virus across the US during the spring wave remains unclear. For the autumn wave, it is challenging to distinguish whether the onset of local outbreaks was driven by infections seeded from other locations through mobility or by resident infections that were already circulating in local communities and may have become more transmissible due to changes in weather or behavior (e.g., changes in humidity or school opening). A further complication with the fall wave is consideration of prior immunity, as a few US locations experienced a relatively large spring wave which would have created a heterogenous landscape of population immunity by the fall. Hence by focusing on the initial seeding event of the A/H1N1pdm influenza virus during spring 2009, we circumvent these issues.

To measure the activity of pandemic influenza, we multiplied the weekly time series of an ILI incidence indicator that represents local ILI visits compiled from electronic medical claims with the positivity rates of A/H1N1pdm influenza in laboratory tests, termed ILI+ (Materials and Methods, Supplementary Information). Using ILI+ time series from March 22^nd^ to July 4^th^, 2009, we inferred transmission links between 220 MSAs with available data. Simulation results confirmed that the inference system was able to reliably reconstruct transmission networks among this subset of MSAs (Supplementary Information, Fig. S9). San Diego, San Antonio, and New York, three locations with early confirmed cases (44, 45), were set as the infection origins for the inference.

The reconstructed transmission network was structurally different from the network for SARS-CoV-2 but exhibited a similar hub-and-spoke pattern (Fig. 3C, Fig. S13). The inferred spatial structure was distinct from the pattern in the fall wave, which started in southeastern US and spread outwards to other regions (28). Fewer high-confidence transmission links were identified for H1N1pdm influenza (Fig. 3D), possibly due to reliance on sparser weekly ILI+ data. Among 220 MSAs, only 168 transmission links were identified, leaving many MSAs without reliably inferred infection sources. Most inferred transmission links occurred within the two weeks prior to April 19^th^, 2009 (Fig. 3D). The estimated transmission network structure was robust to the threshold of occurrence probability (Fig. S14), selected infection origins for the inference (Fig. S15), and the number of initial infections (Fig. S17).

Changes in testing capacity during the early stage of a pandemic can bias estimates of the number of infections of H1N1pdm influenza. For H1N1pdm, testing capacity ramped up quickly in April 2009. On May 3^rd^, 2009, the CDC reported that the majority of confirmed influenza viruses were H1N1pdm (46, 47). During this expansion of testing, using positivity rates of H1N1pdm can partially account for the rapid change in the denominator, though positivity rates will quickly saturate when H1N1pdm became dominant. To address this issue, we combined the syndromic ILI visits data and lab-confirmed positivity rates to quantify the number of H1N1pdm infections; however, the detection rate of H1N1pdm may still have increased over time, particularly during March and April 2009 when tests for H1N1pdm were more limited.

To assess the effect of increasing detection during the early weeks of 2009 pandemic, we performed sensitivity analyses assuming a weekly 20% or 40% increase in the detection rate during the first five weeks of the pandemic, before reaching a plateau in week six. (Note, the observed positivity rate of H1N1pdm sharply increased from week five to week six). Specifically, we modified the ILI+ time series to adjust for this under-detection (i.e., the ILI+ in each MSA in week 𝑡 was multiplied by 1/(80%)^#$𝑡^ or 1/(60%)^#$𝑡^ for 1 ≤ 𝑡 ≤ 5) and re-ran the inference. The resulting transmission networks remained structurally similar except for a few early, long-range transmission pathways (Supplementary Information, Fig. S18). These altered transmission links were originally uncertain with occurrence probabilities close to the selection threshold.

### The role of stochastic processes in early spatial spread

The reconstructed transmission networks revealed shared transmission hubs among the two pandemics, such as New York, Chicago, Seattle, Atlanta, Dallas, and Houston (Fig. 4A). Notably, New York served as a national hub for both pandemics and disseminated virus across long distances via air travel. All these transmission hubs, except Seattle, were among the top 10 MSAs with the largest number of international travelers to other countries in 2009 and 2020 (Table S1). However, other MSAs with large international travel volume, such as Miami, Los Angeles, and San Francisco, were not identified as major disease transmission hubs, suggesting that international travel volume alone is not sufficient to predict the role of MSAs in spatial spread.

**Figure 4.**
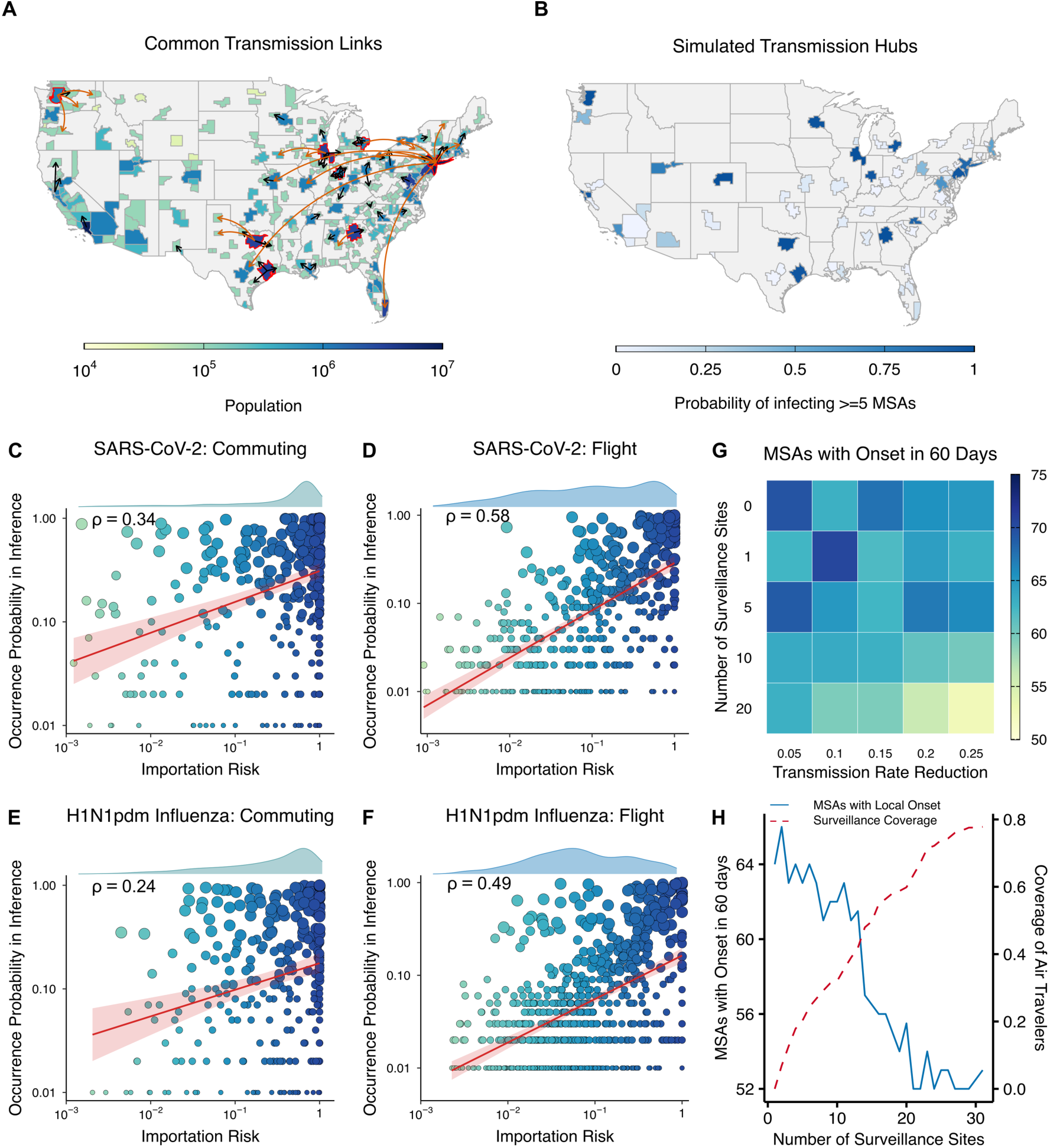
Mobility and early spatial spread. (**A**). The inferred common transmission links for SARS-CoV-2 and H1N1pdm influenza. The color of MSAs represents population size. Red contours highlight the common transmission hubs that infected at least 5 other MSAs in both pandemics. (**B**). Simulated transmission hubs under a variety of scenarios. The color represents the probability of each MSA infecting at least 5 other MSAs among all 980 simulated outbreaks. (C). The Pearson correlation 𝜌 between the importation risk following each commuting link and its occurrence probability in the inference is shown (both the importation risk and occurrent probabilities were log-transformed). Each node represents a commuting link, with node size proportional to its occurrence probability in inference and node color reflecting the importation risk. The red line is a linear regression to the data at the log scale, and the shaded area shows the 95% CI of the fitted values. The distribution of log-transformed importation risk is presented above the panel. (**D**). The same analysis for the importation risk of SARS-CoV-2 through flights. The results for the importation risk of H1N1pdm influenza through commuting and flights are presented in (**E**) and (**F**), respectively. (**G**). The number of MSAs experiencing local onsets within the first 60 days under different surveillance and intervention scenarios for an outbreak originating from New York. The number of surveillance sites and the magnitude of transmission rate reduction were varied. For each scenario, 100 independent simulations were performed. The color shows the median value from 100 outcomes. (**H**). The number of MSAs that had local onsets in the first 60 days with different coverage of wastewater surveillance. The transmission rate reduction was fixed at 25% for interventions. The solid line shows the median values obtained from 100 independent simulations. The dash line shows the cumulative coverage of air travelers.

To model a future respiratory virus pandemic with unknown characteristics, we performed simulations for a variety of outbreak origins, transmissibility, and superspreading potentials (Supplementary Information, Table S2). Consistent with the inference results, the same regional population centers were persistently identified as major infection sources of other MSAs (Fig. 4B), indicating predictable transmission hubs predisposed by mobility patterns.

To examine how well human mobility explains the inferred transmission pathways, we used two mobility-informed heuristics to identify the infection source of each MSA: 1) the volume of incoming mobility, and 2) the infection importation risk (Supplementary Information). For the former, the infection source of an MSA was estimated to be the location contributing to the largest incoming mobility until its onset time. For the latter, we additionally considered the prevalence of infection in other MSAs and computed the expected number of infections imported from each MSA. Mobility volume captured 36.3% (61/168) and 43.1% (131/304) of the inferred transmission links for H1N1pdm influenza and SARS-CoV-2, respectively, whereas infection importation risk captured 34.5% (58/168) and 53.6% (163/304). For both viruses, the infection importation risk for commuting and air travel was positively correlated with the occurrence probabilities of transmission links in the inference (Fig. 4C-F). Air travel better explained inferred transmission pathways than commuting with a stronger correlation for both pandemics; however, the correlations for both mobility modes were only moderate.

To further investigate the predictive power of mobility, we performed inference for the simulated outbreak in Fig. 1A using mobility volume and importation risk. For this idealized outbreak, mobility volume had a precision and recall of 52.5% and 51.7%, while importation risk reached 74.0% and 72.8%, both underperforming the inference algorithm (86.4% and 88.6%). This analysis suggests that stochasticity can render the structure of realized transmission networks less predictable, compromising the inference accuracy of mobility-based heuristics.

### Curtail early spatial spread for future outbreaks

The stochastic nature of spatial spread poses significant challenges for the timely detection and control of future pandemics. Wastewater surveillance at airports offers a practical, cost-effective strategy to detect imported infections (48, 49). We conducted simulations to evaluate the impact of airport wastewater surveillance at key transmission hubs (Fig. 4B), coupled with subsequent reductions in local transmission, on limiting the geographic spread of a hypothetical outbreak originating from New York (Supplementary Information). Specifically, we analyzed the number of MSAs experiencing local onsets within the first 60 days, varying the number of surveillance sites and the magnitude of local transmission reduction. Although simulation outcomes varied widely under each strategy (Fig. S19), expanding surveillance coverage generally slowed spatial spread, provided local transmission was sufficiently reduced (Fig. 4G, Fig. S20). However, effective containment of geographic spread requires broader surveillance beyond the top transmission hubs. For instance, adding surveillance from 5 to 10 sites did not meaningfully improve outcomes, even though the coverage of air travelers increased from 24.3% to 35.6% (Fig. 4H). The performance was stabilized until 20 hubs were selected (covering 63.5% of air travelers), suggesting that stochastic importation events require sufficient surveillance coverage to effectively curb early spatial spread. Similar findings were obtained for another outbreak originating from Minnesota (Fig. 1A, Fig. S21). These simulations suggest that coordinated wastewater surveillance across multiple key locations can potentially inform early containment of future outbreaks.

## Discussion

The early cryptic transmission of emerging respiratory viruses is difficult to directly observe due to limited surveillance data. Here, we inferred the transmission networks of SARS-CoV-2 and H1N1pdm influenza using high-resolution disease data and a new inference framework. Our inference revealed that both viruses reached most MSAs within a span of weeks, leaving a short time window for decision-making and effective interventions. The rapid geographic dissemination underscores the challenges of early-stage containment of pandemic respiratory viruses.

Beyond reconstructing historical spread of the last two pandemics, our study also provides a generalizable framework to infer early epidemic dynamics that may be applied to other pathogens. By combining mechanistic modeling, mobility integration, and ensemble-based inference, this approach provides a robust, data-efficient method to resolve spatial transmission patterns in the critical early window.

Our inference approach highlights the important role of mobility, particularly air travel, in driving the geographic expansion of novel pathogens. However, other factors such as school schedules (50), winter holidays (51), meteorological conditions (52, 53), and reactive human behaviors (54–57) can also influence transmission dynamics. Using mobility data alone to predict the spatial spread of pathogens within the US, where MSAs are highly connected through multiple pathways, remains challenging. Given these inherent uncertainties, accurately reconstructing transmission routes across locations may require multiple data sources. Genomic data could provide additional evidence to refine the inference of spatial spread (20–23), although sequencing is typically scarce in the early stage of a new outbreak. Expanding the coverage and reducing the bias of genomic data collection (58) may support improved inference using multimodal data streams. New inference frameworks that jointly use information from mobility and genomic data can be developed in future studies (e.g., modifying a model-less likelihood-based approach to reconstruct transmission chains (59–61)).

While many modeling studies demonstrated the potential benefits of wastewater surveillance, barriers still exist in its real-world implementation such as coordination among different stakeholders and financial support. Modeling studies can focus on the comparison of wastewater and other modes of surveillance. For instance, future studies can incorporate realistic reporting delay and sampling strategies for clinical cases and quantify the improvement of using wastewater over clinical cases in early outbreak detection.

The spatiotemporal pattern of infections may shift as pandemic respiratory pathogens transition to endemicity. This shift may be influenced by the evolving immunological landscape, demographic structure, viral evolution, vaccination coverage, and long-term changes in human behaviors.

Understanding and anticipating the equilibrium spatiotemporal dynamics can inform improved response efforts to reduce disease burdens during the endemic phase. For novel pathogens for which there is partial protection from existing vaccines, differential vaccination coverage would affect population susceptibility (62), producing an uneven immunological landscape that would impact the spatiotemporal transmission dynamics and should be evaluated in future studies.

There are a few limitations in this study. As the model does not include detailed social structure, simulations cannot represent the heterogeneous transmission dynamics across age-groups and communities within each MSA. Additionally, we did not include international travels thus may miss some introductions from other countries. Further, the simulated surveillance and intervention strategies are a proof-of-concept and may not be practical in real-world settings. Studies on more realistic approaches are needed to guide actionable responses in future works.

## Materials and Methods

### Data

**H1N1pdm influenza.** We used electronic medical claims data compiled by IMS Health to create weekly influenza-like illness (ILI) time series in 2009 for 336 US cities (35). IMS Health data covered 62% of US outpatient visits in 2009. The IMS Health medical claims data were de-identified and aggregated at the weekly and city level prior to sharing with our research team. To measure ILI incidence in a location during a given week, we used the ILI incidence indicator for location 𝑖 in week 𝑡, defined as

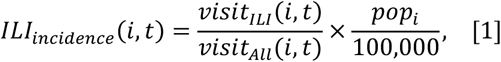

where 𝑣𝑖𝑠𝑖𝑡_𝐼𝐿𝐼_(𝑖, 𝑡) is the number of ILI visits, 𝑣𝑖𝑠𝑖𝑡_𝐴𝑙𝑙_(𝑖, 𝑡) is the total number of medical visits, and 𝑝𝑜𝑝_𝑖_ is the population of location 𝑖. We aggregated the ILI incidence indicator data to the MSA scale and obtained weekly 𝐼𝐿𝐼_𝑖𝑛𝑐𝑖𝑑𝑒𝑛𝑐𝑒_ time series for 220 MSAs, covering 75.5% of the US population in 2009. To better capture the activity of H1N1pdm influenza, we further used the regional positivity rate of influenza A (H1N1)pdm09 in laboratory tests reported to the US CDC by public health laboratories (63). We multiplied the weekly time series of ILI incidence indicator in each MSA with the concurrent positivity rate in its corresponding HHS region, termed ILI+ incidence (64), to track the activity of H1N1pdm influenza. In the inference, we used weekly ILI+ incidence time series from March 22^nd^, 2009 to July 4^th^, 2009.

**SARS-CoV-2.** Due to limited surveillance capacity and asymptomatic and presymptomatic transmission, COVID-19 infections in the US were severely underreported. We used estimated daily new infections (both reported and unreported) in 3,142 US counties in 2020 (31). This estimation employed a dynamic data-driven transmission model that accounted for mobility across US counties, heterogeneous transmission rates, time-varying infection ascertainment rates, and reporting delay from infection acquisition to case confirmation. The estimated total infection numbers were validated independently by serological data collected at different locations and time points (31). The estimated daily new infections were aggregated to the daily infections for the 375 MSAs, accounting for 83.8% of the US population in 2020. We used daily infection data from February 21^st^, 2020 to May 10^th^, 2020 for the inference.

**Human mobility.** The county-to-county commuting data were compiled from American Community Survey publicly available at the US Census Bureau (65). Inter-MSA commuting data were obtained by aggregating county-level commuting data to the MSA level. For SARS-CoV-2, inter-MSA commuting after March 1^st^, 2020 was further adjusted using the relative change of inter-county movement measured by mobile phone-derived foot-traffic data (66) (Supplementary Information). The air travel data were downloaded from the US Bureau of Transportation Statistics (67) and recorded the monthly number of passengers on flights between US airports. We evenly distributed the monthly passenger numbers to each day and aggregated the daily passenger numbers to the MSA level based on the location of airports.

### Transmission model

To simulate the spread of novel respiratory viruses, we developed a stochastic data-driven metapopulation model informed by commuting and flight data across MSAs in the US. Transmission dynamics are described by the following equations for a metapopulation SIRS model.

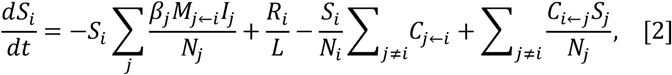

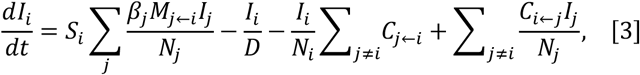

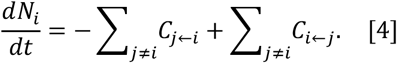

Here 𝐷 is the average infectious period, 𝐿 is the mean duration of immunity, 𝑀_j➛𝑖_ is the fraction of population living in location 𝑖 commuting to location 𝑗, and 𝐶_j➛𝑖_ is the number of passengers flying from location 𝑖 to location 𝑗. The transmission model is integrated stochastically. Specifically, we used a negative binomial distribution to generate the number of secondary infections, representing the superspreading potential of the pathogen. Population migration across locations via air travel was modeled using a multinomial distribution. During model integration, we tracked the number of infections in each location attributed to commuting and air travel. As a result, we can define transmission by specific mobility modes (see details in Supplementary Information). Similar data-driven model construct has been used in our previous study to simulate spatial spread of respiratory pathogens (68).

### Definition of spatial spread

We used a piecewise linear fitting to identify the onset time in each location. This approach has been used in previous studies on spatial spread of influenza (30, 38). For each MSA 𝑖, we used a bi-linear fitting to identify the onset time 𝑡_𝑜𝑛𝑠𝑒𝑡_ of this location. We computed the cumulative number of infections attributed to other locations (through commuting and air travel) until time 𝑡_𝑜𝑛𝑠𝑒𝑡_ and selected the location 𝑗 that contributed most seeds to location 𝑖 as the source MSA. The transmission link 𝑗 → 𝑖 is defined as a ground-truth transmission from 𝑗 to 𝑖. As we can track the number of infections caused by commuting and air travel, the ground-truth transmission link 𝑗 → 𝑖 was defined as the mobility mode that contributed most infections (Supplementary Information).

### Inference framework

In the inference, we first rank all locations based on the estimated local onset time. Starting from the first location, we sequentially estimated the most likely infection source of each location that best predicted the number of infections after onset in that location. To do this, we tested all possible candidate models with mobility from each potential infection source, generated forecasts using each model at the onset week, and selected the model with the lowest forecast error. If none of the candidate models outperformed the null model (i.e., no mobility linked to the focal location, thus producing zero infection), we deemed that no reliable infection source was found for this location in this round of inference. For these locations, we changed their ranking of onset time and performed a few additional rounds of inference, assuming the estimated onset week may not be accurate. Once a transmission link was identified, we added the mobility between the inferred infection source and the focal location to the transmission model, which was used as the null model in the next round of inference. This sequential procedure was performed until all locations were examined in the inference. During the inference, the identified mobility links responsible for cross-location transmission were gradually added to the underlying transmission model. This process essentially reconstructed the minimal “backbone” transmission network that best explained the spatial spread of the pathogen. The inference framework has a few designs to address uncertainties in onset time estimation and the setting of seeding locations. A pseudo- code of the inference algorithm is provided in Supplementary Information.

As the transmission model has stochastic dynamics, different realizations of the inference algorithm may result in different transmission networks. Performing independent realizations of inference provide a means of quantifying the uncertainty of the inferred transmission links. For each analysis, we performed 100 independent realizations of inference and formed an ensemble of reconstructed transmission networks. To select high-confidence transmission links, we used a threshold for the occurrence probability in the ensemble to reconstruct the transmission network. The threshold was optimized using F1-score to balance the precision and recall in the inference results.

Extensive sensitivity analyses were performed to examine the robustness of the inference algorithm to the mis-selection of seeding locations, transmission model misspecification, and data missingness. Experiments on synthetic outbreaks show that the inference can still yield satisfactory results even when these issues exist in model configurations and data quality (Supplementary Information).

### Inference using real-world data

In the inference for SARS-CoV-2 and H1N1pdm influenza, we mapped the observed disease data to the number of new infections generated by the transmission models (see details in Supplementary Information). In the transmission models, we used epidemiological parameters for each pathogen reported in previous studies (69–73) and adjusted location-specific transmission rates to generate forecasts. The seeding locations of SARS-CoV-2 were selected as Seattle and New York, informed by the estimated onset times and genomic studies (20). For H1N1pdm influenza, seeding locations (San Diego, San Antonio, and New York) were selected based on early case reports of infections (44, 45). We estimated the rough number of initial infections in seeding locations and found that inference results were robust to the choice of seeding locations and range of initial infections (Supplementary Information).

### Modeling wastewater surveillance at transmission hubs

We performed simulations to assess the effects of wastewater surveillance among air travelers and subsequent interventions to slow down the spatial spread of a novel pathogen. Since the cryptic transmission in the outbreak origin is difficult to detect, we deployed wastewater surveillance in transmission hubs identified in Fig. 4B, agnostic of the origin of the novel pathogen. Specifically, we selected a given number of transmission hubs following a decreasing order of the probabilities of each MSA infecting ≥ 5 other MSAs among the simulations in Fig. 4B. Previous research estimated that the detection rate of an infected individual through wastewater surveillance on flights is in the range of 11-22% for SARS-CoV-2 (49). In our simulations, we used a 20% detection rate for each infected individual among air travelers arriving at the MSAs with wastewater surveillance. Interventions were enacted when at least one infection was detected using wastewater surveillance, or the cumulative local infections in an MSA reached a threshold (e.g., some locally infected people may seek medical treatment and be discovered at hospitals), whichever came first. We assume the interventions will reduce the local transmission rate by a certain percentage due to risk aversion, social distancing, or other control policies. Simulations were performed for outbreaks originating from New York and Minnesota, with transmission characteristics similar to H1N1pdm influenza (Supplementary Information).

## Data Availability

Data used in the study are available online at https://github.com/SenPei-CU/PandemicInference.

## Acknowledgments

This study was supported by funding from National Natural Science Foundation of China 12371516 (RZ), National Science Foundation DMS-2229605 (SP), National Institute of General Medical Sciences R35GM156799 (SP), Centers for Disease Control and Prevention U01CK000592 (JS, SP) and 75D30122C14289 (JS), National Institute of Allergy and Infectious Diseases R01AI163023 (JS), Princeton Catalysis Initiative (BTG), Princeton Precision Health (BTG), and High Meadows Environmental Institute (BTG). The findings and conclusions in this report are those of the authors and do not necessarily represent the official position of the US National Institutes of Health, Centers for Disease Control and Prevention, or Department of Health and Human Services.

## Author Contributions

SP conceived the study, RZ, RD, SL, and SP performed the analysis, RD, SL, CV and SP curated data, RZ, JS, BTG, CV, and SP contributed methodologies, RZ, RD, SL, QY, JS, BTG, CV and SP investigated the results, RZ, RD, SL, and SP drafted the original manuscript, QY, JS, BTG, and CV revised and reviewed the manuscript.

