## Supplementary Information for "Reconstructing the early spatial spread of pandemic respiratory viruses in the United States"

### Supporting Information for
Reconstructing the early spatial spread of pandemic respiratory viruses in the United States

Renquan Zhang[1], Rui Deng[1†], Sitong Liu[1†], Qing Yao[2], Jeffrey Shaman[2,3], Bryan T Grenfell[4], Cécile Viboud[5], Sen Pei[2*]

[1]School of Mathematical Sciences, Dalian University of Technology, Dalian, China

[2]Department of Environmental Health Sciences, Mailman School of Public Health, Columbia University, New York, NY, USA

[3]Columbia Climate School, Columbia University, New York, NY, USA

[4]Department of Ecology and Evolutionary Biology, Princeton University, Princeton, NJ, USA

[5]Fogarty International Center, National Institutes of Health, Bethesda, MD, USA

*Corresponding author: Sen Pei.

†These authors contributed equally to this work.

#### Data

**SARS-CoV-2.** Due to limited surveillance capacity and asymptomatic and presymptomatic transmission, COVID-19 infections in the US were severely underreported. To address this issue, instead of using the daily confirmed COVID-19 cases, we used estimated daily new infections (both reported and unreported) in 3,142 US counties, as reported in Ref. (1). This estimation employed a dynamic data-driven transmission model that accounted for mobility across US counties, heterogeneous transmission rates, time-varying infection ascertainment rates, and reporting delay from infection acquisition to case confirmation. The estimated total infection numbers were validated independently by serological data collected at different locations and time points (1). The estimated daily new infections were smoothed using a 7-day moving average and aggregated to the daily infections for the 375 MSAs defined in 2020 (note the designation of MSAs changed over time). The 375 MSAs accounts for 83.8% of the US population in 2020. We used daily infection data from February 21st, 2020 to May 10th, 2020 for the inference. The national time series of estimated daily new infections is shown in Fig. S1A.

**H1N1pdm influenza.** We used electronic medical claims data compiled by IMS Health to create weekly influenza-like illness (ILI) time series in 2009 for 336 US cities (2). IMS Health data covered 62% of US outpatient visits in 2009. To measure ILI incidence in a location during a given week, as in Ref. (2), we used the ILI incidence indicator for location $i$ in week $t$, defined as

$$ILI_{incidence}(i,t) = \frac{visit_{ILI}(i,t)}{visit_{All}(i,t)} \times \frac{pop_i}{100{,}000}, \quad [1]$$

where $visit_{ILI}(i,t)$ is the number of ILI visits, $visit_{All}(i,t)$ is the total number of medical visits, and $pop_i$ is the population of location $i$. This metric was found highly synchronous with the influenza surveillance data from the US CDC (2) and has been used in many previous studies on the spatial spread of influenza (3–5). We aggregated the ILI incidence indicator data to the MSA scale and obtained weekly $ILI_{incidence}$ time series for 220 MSAs, covering 75.5% of the US population in 2009. To better capture the activity of H1N1pdm influenza, we further used the regional positivity rate of influenza A (H1N1)pdm09 in laboratory tests reported to the US CDC by public health laboratories (6). We multiplied the weekly time series of ILI incidence indicator in each MSA with the concurrent positivity rate in its corresponding HHS region, termed ILI+

incidence (7), to track the activity of H1N1pdm influenza (i.e., $ILI+_{incidence}(i,t) = ILI_{incidence}(i,t) \times pr(h,t)$, where $pr(h,t)$ is the positivity rate for HHS region $h$ in week $t$). Since laboratory tests for A (H1N1)pdm09 were performed beginning the week of April 19th, 2009, the weekly ILI+ incidence time series was truncated due to the lack of positivity rate data before that date. To impute ILI+ incidence data prior to April 19th, 2009, we fitted an exponential growth curve to the data from the four weeks after April 19th, 2009 and estimated ILI+ incidence in the four weeks prior to April 19th, 2009. In the inference, we used weekly ILI+ incidence time series from March 22nd, 2009 to July 4th, 2009. The national weekly ILI+ incidence time series is shown in Fig. S1B.

**Human mobility.** The county-to-county commuting data for the 2009 H1N1 pandemic were compiled from the 2009-2013 5-Year American Community Survey (ACS) publicly available at the US Census Bureau (8). Inter-MSA commuting data were obtained by aggregating county-level commuting data to the MSA level. For SARS-CoV-2, we used the county-to-county commuting data from the 2016-2020 5-Year ACS Commuting Flows (9) and aggregated the data to the MSA scale. Prior to March 1st, 2020, we used the commuting data from ACS to prescribe the inter-MSA commuting. After March 1st, 2020, the census survey data were no longer representative due to changes in mobility behavior following implementation of non-pharmaceutical interventions. We therefore used estimates of the reduction of inter-county visitors to points of interest (POI) (e.g. restaurants, stores, etc.) from SafeGraph (10) to account for the change of inter-county movement on a county-by-county basis. For instance, if the number of inter-county visitors to a county was reduced by 10% on a given day relative to the baseline on March 1st, the number of commuters to this county would be reduced by 10% accordingly. The reduced inter-county mobility was then aggregated to the MSA level. This approach has been used in our previous studies (1, 11). The airline travel data were downloaded from the US Bureau of Transportation Statistics (12) and recorded the monthly number of passengers on flights between US airports. We evenly distributed the monthly passenger numbers to each day and aggregated the daily passenger numbers to the MSA level based on the location of airports. The commuting and airline travel data are shown in Fig. S2.

**Transmission model**

To simulate the spread of a hypothetical novel respiratory virus, we developed a stochastic data-driven metapopulation model informed by commuting and flight data for all 367 MSAs (in 2009) in the US. An MSA is a geographical region with a relatively high population density at its urban center (or centers) and close social and economic ties throughout the region, measured by commuting and employment. As MSA is not defined by administrative boundaries (cities, counties, or states), it serves as a natural geographical unit to characterize the spatial spread of respiratory diseases because of the frequent population mixing within MSAs. The total population in the 367 MSAs was 261.1M in 2009, accounting for 85.1% of US population (306.8M) at that time.

Transmission dynamics are described by the following equations for a metapopulation SIRS model.

$$\frac{dS_i}{dt} = -S_i \sum_j \frac{\beta_j M_{j\leftarrow i} I_j}{N_j} + \frac{R_i}{L} - \frac{S_i}{N_i} \sum_{j\neq i} C_{j\leftarrow i} + \sum_{j\neq i} \frac{C_{i\leftarrow j} S_j}{N_j}, \quad [2]$$

$$\frac{dI_i}{dt} = S_i \sum_j \frac{\beta_j M_{j\leftarrow i} I_j}{N_j} - \frac{I_i}{D} - \frac{I_i}{N_i} \sum_{j\neq i} C_{j\leftarrow i} + \sum_{j\neq i} \frac{C_{i\leftarrow j} I_j}{N_j}, \quad [3]$$

$$\frac{dN_i}{dt} = -\sum_{j\neq i} C_{j\leftarrow i} + \sum_{j\neq i} C_{i\leftarrow j}. \quad [4]$$

Here $D$ is the average infectious period, $L$ is the mean duration of immunity, $M_{j\leftarrow i}$ is the **fraction** of population living in location $i$ commuting to location $j$, and $C_{j\leftarrow i}$ is the **number** of passengers flying from location $i$ to location $j$. The commuting matrix $\{M_{j\leftarrow i}\}$ is static over time for H1N1pdm influenza and is adjusted by mobile phone-derived POI visit data on a daily basis for SARS-CoV-

2. The flight matrix $\{C_{j\leftarrow i}\}$ changes over time based on real-world flight data and is asymmetric between MSAs. The population in each MSA is updated using Eq. [4]. The commuting data capture recurrent short-range human mobility for work (mostly ground transportations –cars, buses, trains, etc.) and the flight data reflect long-range human mobility across MSAs. To reduce the number of mobility links in the transmission model, we removed commuting links with less than 200 visitors per day and airline travel links with less than 100 visitors per day. The removed mobility links only accounted for 0.53% of the total travel volume. Therefore, we do not expect this removal substantially impacted simulation outcomes.

The first term on the right-hand side of Eq. [2] represents the transmission caused by commuting. Specifically, $S_i M_{j\leftarrow i}$ susceptible individuals living in location $i$ will mix with people in location $j$, for whom the probability of being infectious is $I_j/N_j$. The parameter $\beta_j$ (termed transmission rate) represents the product of the number of contacts and the probability of infection upon a contact with an infectious individual in location $j$. The latter two terms on the right-hand side of Eq. [2] represent population migration between MSAs through airline travel. The number of passengers moving from $i$ to $j$ is $C_{j\leftarrow i}$, among which $S_i/N_i$ fraction is susceptible people in location $i$. The term $S_i/N_i\ \sum_{j\neq i} C_{j\leftarrow i}$ is the total number of susceptible people moving out of location $i$. Similarly, the total number of infectious people moving into location $i$ is $\sum_{j\neq i} C_{i\leftarrow j} S_j/N_j$.

**Modeling superspreading and stochastic dynamics**

**Superspreading.** Many respiratory diseases exhibited individual variation in disease transmission, leading to superspreading events in which a small number of individuals infected disproportionately large numbers of secondary cases (13). Superspreading events have been extensively documented for a range of pathogens, including SARS-CoV (14–17), measles (18, 19), MERS-CoV (20, 21), and more recently, SARS-CoV-2 (22, 23). For pathogens with a strong superspreading potential, the establishment of sustained transmission may require multiple introductions of infections as most infected individuals can only infect a limited number of secondary cases and may die out spontaneously (13). As a result, the superspreading potential of a pathogen can introduce stochasticity to the early spatial spread of the pathogen.

**Negative binomial distributions.** To model superspreading, we used a negative binomial (NB) distribution to generate the number of secondary infections. The NB distribution is widely used to represent individual heterogeneity in disease transmission (13). Here, we parameterized the NB distribution using the mean value $m$ and the dispersion parameter $r$. Specifically, for a random variable $z{\sim}NB(m,r)$, the probability density function is:

$$\Pr(z=k)=\frac{\Gamma(r+k)}{k!\,\Gamma(r)}\left(\frac{r}{r+m}\right)^k\left(\frac{m}{r+m}\right)^r, \text{for } k=0,1,2,\ldots \quad [5]$$

Note that the variance of the NB distribution is:

$$\sigma^2=m+\frac{m^2}{r}. \quad [6]$$

The dispersion parameter $r$ controls the variance of the NB distribution. For $r\to\infty$, $\sigma^2\to m$, the NB distribution converges to a Poisson distribution. For $r\to 0$, $\sigma^2\to\infty$, the variance of the NB distribution diverges. For infectious diseases, a smaller dispersion parameter $r$ indicates a larger heterogeneity in disease transmission, thus a stronger superspreading potential. A list of estimated dispersion parameters for several infectious diseases can be found in Ref. (13). Suppose each infectious individual $i$ can generate $z_i$ new infections, where $z_i{\sim}NB(m,r)$. Using the moment generating function, it can be shown that the total number of new infections generated by $n$ infectious individuals is $Z_n=\sum_{i=1}^{n} z_i\ {\sim}NB(nm,nr)$ (24). The coefficient of variation, defined as the ratio between the standard deviation $\sigma_{Z_n}$ to the mean value $\mu_{Z_n}=nm$, is

$$\frac{\sigma_{Z_n}}{\mu_{Z_n}}=\sqrt{\frac{1}{nm}+\frac{1}{nr}}=\sqrt{\frac{1}{\mu_{Z_n}}+\frac{1}{nr}}. \quad [7]$$

This equation indicates that when the number of infectious individuals, $n$, increases, the relative variation of new infections becomes smaller. In other words, the role of individual variation diminishes when there are more infections in the population.

**New infections attributed to commuting.** We integrated the transmission model in Eqs. [2-4] stochastically on a daily time step. We first generate the number of new infections in location $i$ acquired in location $j$ due to commuting. In the transmission model, the number of infectious individuals in location $j$ is $I_j$. On average, each will infect $\beta_j \sum_k S_k M_{j\leftarrow k} / N_j$ susceptible individuals present in location $j$ (coming from all locations). The total new infection each day can be drawn from

$$\Delta I_j \sim NB\left(I_j \beta_j \sum_k S_k M_{j\leftarrow k} / N_j, I_j r\right). \quad [8]$$

Among those new infections, we use a multinomial distribution to distribute them to different locations:

$$\left(Seed^1_{1\leftarrow j}, \cdots, Seed^1_{L\leftarrow j}\right) \sim Multinomial\left(\Delta I_j; \frac{S_1 M_{j\leftarrow 1}}{\sum_k S_k M_{j\leftarrow k}}, \cdots, \frac{S_n M_{j\leftarrow L}}{\sum_k S_k M_{j\leftarrow k}}\right). \quad [9]$$

The number of infections in location $j$ caused by infections in location $i$ is $Seed^1_{i\leftarrow j}$. This calculation is compatible with the dynamics in Eqs. [2-3].

**Imported infections attributed to airline travel.** The average number of infectious individuals in location $i$ migrating from location $j$ can be generated using a multinomial distribution as well. Note, we define the population moving from location $j$ to the same location $j$ as $C_{j\leftarrow j} = N_j - \sum_{k\neq j} C_{j\leftarrow k}$ (including the population staying in location $j$).

$$\left(Seed^2_{1\leftarrow j}, \cdots, Seed^2_{L\leftarrow j}\right) \sim Multinomial\left(I_j; \frac{C_{1\leftarrow j}}{\sum_k C_{k\leftarrow j}}, \cdots, \frac{C_{L\leftarrow j}}{\sum_k C_{k\leftarrow j}}\right). \quad [10]$$

The number of infections in location $j$ migrated from location $i$ is $Seed^2_{i\leftarrow j}$.

**The total number of infections** in location $i$ seeded from location $j$ is

$$Seed_{i\leftarrow j} = Seed^1_{i\leftarrow j} + Seed^2_{i\leftarrow j}. \quad [11]$$

Note the model can distinguish the number of new infections attributed to flight and commuting. As a result, we can define transmission by specific mobility modes.

**Defining spatial spread**

**Estimating the onset time of local transmission.** We used a piecewise linear fitting to identify the onset time in each location. This approach has been used in previous studies on spatial spread of influenza (3, 5). We fitted a bi-linear trend to the time series of disease data. The breakpoint of the linear fitting was used to define the onset time of local transmission. For SARS-CoV-2, the fitting was performed for the daily time series from the beginning of the study period (February 21st, 2020) to the date reaching the cumulative infections of 1000. If the cumulative infections did not reach 1000 in the first 80 days, we used the data until the first peak in the 80-day period. For H1N1pdm influenza, we used weekly ILI+ time series from March 22nd, 2009 to the week reaching the cumulative ILI+ incidence of 0.2. We show the examples of estimating the onset times for SARS-CoV-2 and H1N1pdm influenza in the San Antonio–New Braunfels, TX MSA in Fig. S3.

**Defining ground-truth transmission links across locations.** For pathogens with superspreading potential, the establishment of local transmission in a new location typically requires multiple seeds introduced from other locations. As the transmission model does not track the secondary infections generated by each individual and there could be many imported infections contributing to local transmission, it is not straightforward to definitively define the infection source of a location. Here, we used the heuristic approach described below.

For each MSA $i$, we used a bi-linear fitting to identify the onset time $t_{onset}$ of this location. We computed the cumulative number of infections attributed to other locations (through commuting and airline travel) until time $t_{onset}$ and selected the location $j$ that contributed most seeds to location $i$ as the source MSA. The transmission link $j \rightarrow i$ is defined as a ground-truth transmission from $j$ to $i$. As we can track the number of infections caused by commuting and airline travel, the ground-truth transmission link $j \rightarrow i$ was defined as the mobility mode that contributed most infections. In the inference, we deemed the result for MSA $i$ was accurate if both the infection source and mobility mode were correctly inferred.

**Simulation of a hypothetical novel respiratory virus**

We simulated the spatial spread of a hypothetical novel respiratory virus. In the simulation, we set model parameters as following: a uniform transmission rate $\beta = 0.375$ for all MSAs, a mean infectious period $D = 4$ days, a mean immunity duration $L = 1.5$ years, and a dispersion parameter $r = 2.36$. The pathogen had a basic reproductive number $R_0 \approx \beta D = 1.5$ in each MSA (25) and the superspreading potential similar to H1N1pdm influenza (26). In the simulation, we used the mobility data (commuting and airline travel) from March 22nd, 2009 to November 21st, 2009, running the simulation for 245 days. 20 infectious people were seeded to initialize an outbreak in Minneapolis–St. Paul–Bloomington, MN–WI MSA, the largest MSA in Minnesota. We selected Minnesota as a hypothetical outbreak origin as Minnesota reported many confirmed detections of H5N1 highly pathogenic avian influenza (HPAI) in commercial and backyard flocks during the 2022 to 2024 period (27). The model was integrated daily and generated the weekly new infections in each MSA, per the reporting cadence of influenza.

To examine the uncertainty of the early spatial spread simulated by the transmission model, we performed 100 independent realizations of the simulation. We recorded transmission links that occurred at least once in these simulations and computed their occurrence probabilities. These transmission links were grouped into five categories based on the occurrence probability ($(0\%, 20\%]$, $(20\%, 40\%]$, $(40\%, 60\%]$, $(60\%, 80\%]$, $(80\%, 100\%]$). We examined the number of transmission links belonging to each category among all transmission links that appeared in the 100 simulations (Fig. 1B). We further showed the distribution of transmission links across the five categories for the transmission links obtained from a single simulation (Fig. 1C).

For each MSA, we identified the onset time of local transmission and infection source produced by each simulation. The entropy of onset time for an MSA $i$ was defined as

$$H\left(t_{onset}^{i}\right) = -\sum_{k} p\left(t_{onset,k}^{i}\right) \log\left(p\left(t_{onset,k}^{i}\right)\right). \quad [12]$$

Here $t_{onset}^{i}$ is the random variable representing the onset date for MSA $i$, $\{t_{onset,k}^{i}\}$ is the set of distinct onset dates for MSA $i$ generated from simulations, $p\left(t_{onset,k}^{i}\right)$ is the probability for $t_{onset,k}^{i}$ in the simulations, and the base of the logarithm is $e$. Similarly, we defined the entropy of infection source as

$$H(O^{i}) = -\sum_{k} p\left(O_{k}^{i}\right) \log\left(p\left(O_{k}^{i}\right)\right). \quad [13]$$

Here $O^{i}$ is the random variable of infection source for MSA $i$, $\{O_{k}^{i}\}$ is the set of unique infection sources for MSA $i$ in simulated outcomes, and $p(O_{k}^{i})$ is the probability for $O_{k}^{i}$ in simulations.

We visualized the transmission network generated from one simulation from different perspectives. Transmission links across MSAs at different layers from the origin MSA showed the hierarchical spatial structure of the transmission network (Fig. S4). Transmission links occurring in different weeks presented the temporal dynamics of the geographical expansion of the pathogen (Fig. S5).

To examine how virus transmissibility and superspreading potential impact the uncertainty of realized transmission links, we performed additional simulations using a wide range of parameter values for the transmission rate $\beta$ and the dispersion parameter $r$. The basic reproductive number in these simulations ranged from 1.6 to 4. For each parameter combination, 100 independent simulations were performed with settings same to those in Fig. 1A. The fraction of stable transmission links that appeared in at least 80% of simulations is shown in Fig. S6. The fraction of stable transmission links varied from 0.6 to 0.78. A less transmissible virus (a low basic reproductive number) and a higher superspreading potential (a low dispersion parameter) tend to generate more variable transmission networks in independent simulations.

**Inference algorithm**

**Background.** Many approaches exist to detect dynamical interactions between time series, such as Granger Causality (28), transfer entropy (29), and convergent cross mapping (30). However, these techniques were designed for general time series and were found to have high false-positive rates in a range of dynamical models (31, 32). Here, we developed a prediction-based inference framework tailored to the spatial spread dynamics of infectious diseases. The principle of predictability improvement is fundamental to many inference techniques. For instance, the Granger Causality uses a vector autoregressive (VAR) model to determine whether including information from one time series can improve the prediction of the other time series. As the underlining VAR model may not well represent the data generation process of observations, this approach may fail in many applications. Changing the VAR model to a more flexible state-space reconstruction model (e.g., the method of analogs) substantially improved the performance of inference in several chaotic dynamical models (31). But to ensure a decent prediction accuracy, this approach requires a large amount of training data, which is typically not available in disease surveillance.

To overcome the challenges of model misspecification and data sparsity, we used a data-driven disease transmission model with spatial structures to incorporate prior knowledge on the connectivity between locations and the nonlinear, stochastic transmission dynamics of pathogens. This explicit representation of key processes in the spatial spread of infectious diseases allows the inference system to identify essential transmission links that can better explain the observed disease data. This data-driven approach may better constrain plausible time series and geographical spread and thus require less data to achieve reliable inference performance.

**Summary of the inference framework.** In the inference, we first rank all locations based on the estimated local onset time. Starting from the first location, we sequentially estimated the most likely infection source of each location that best predicted the number of infections after onset in that location. To do this, we tested all possible candidate models with mobility from each potential infection source, generated forecasts using each model at the onset week, and selected the model with the lowest forecast error. If none of the candidate models outperformed the null model (i.e., no mobility linked to the focal location, thus producing zero infection), we deemed that no reliable infection source was found for this location in this round of inference. For these locations, we changed their ranking of onset time and performed a few additional rounds of inference, assuming the estimated onset week may not be accurate. Once a transmission link was identified, we added the mobility between the inferred infection source and the focal location to the transmission model, which was used as the null model in the next round of inference. This sequential procedure was performed until all locations were examined in the inference. During the inference, the identified mobility links responsible for cross-location transmission were gradually added to the underlying transmission model. This process essentially reconstructed the minimal "backbone" transmission network that best explained the spatial spread of the pathogen. An example of the inference for three locations is provided in Fig. S6.

**Efficient epidemic forecasting.** Since a large number of forecasts needs to be generated using a high-dimensional dynamical model, an efficient forecasting technique is needed. Here, we coupled the transmission model with an efficient data assimilation algorithm, the ensemble adjustment Kalman filter (EAKF) (33), to generate probabilistic predictions (34). The EAKF assumes a Gaussian distribution of both the prior and likelihood and adjusts the prior distribution to a posterior using Bayes' rule deterministically. To represent the state-space distribution, the EAKF maintains an ensemble of system state vectors acting as samples from the distribution. In particular, the EAKF assumes that both the prior distribution and likelihood are Gaussian and thus can be fully characterized by their first two moments (mean and variance). The update scheme for ensemble members is computed using Bayes' rule (posterior ∝ prior × likelihood) via the convolution of the two Gaussian distributions. For observed state variables (e.g., daily new infections), the posterior of the $i$th ensemble member is updated through

$$y_{t,post}^{i} = \frac{\sigma_{t,obs}^{2}}{\sigma_{t,obs}^{2} + \sigma_{t,prior}^{2}} \bar{y}_{t,prior} + \frac{\sigma_{t,prior}^{2}}{\sigma_{t,obs}^{2} + \sigma_{t,prior}^{2}} y_{t}^{o} + \sqrt{\frac{\sigma_{t,obs}^{2}}{\sigma_{t,obs}^{2} + \sigma_{t,prior}^{2}}} \left(y_{t,prior}^{i} - \bar{y}_{t,prior}\right). \quad [14]$$

Here $y_{t,post}^{i}$ and $y_{t,prior}^{i}$ are the posterior and prior of the observed variable for the $i$th ensemble member at time $t$; $\bar{y}_{t,prior}$ is the mean of the prior observed variable; $\sigma_{t,obs}^{2}$ and $\sigma_{t,prior}^{2}$ are the variances of the observation and the prior observed variable; and $y_{t}^{o}$ is the observation at time $t$. Unobserved variables (e.g., susceptible population) and parameters (e.g., transmission rate) are updated through their covariability with the observed variable, which can be computed directly from the ensemble. In particular, the $i$th ensemble member of unobserved variable or parameter $x^{i}$ is updated by

$$x_{t,post}^{i} = x_{t,prior}^{i} + \frac{\sigma\left(\left\{x_{t,prior}\right\}_{n}, \left\{y_{t,prior}\right\}_{n}\right)}{\sigma_{t,prior}^{2}} \left(y_{t,post}^{i} - y_{t,prior}^{i}\right). \quad [15]$$

Here $x_{t,post}^{i}$ and $x_{t,prior}^{i}$ are the posterior and prior of the unobserved variable or parameter for the $i$th ensemble member at time $t$; and $\sigma\left(\left\{x_{t,prior}\right\}_{n}, \left\{y_{t,prior}\right\}_{n}\right)$ is the covariance between the prior of the unobserved variable or parameter $\left\{x_{t,prior}\right\}_{n}$ and the prior of the observed variable $\left\{y_{t,prior}\right\}_{n}$ at time $t$. In the EAKF, variables and parameters are updated deterministically such that the higher moments of the prior distribution are preserved in the posterior.

For the EAKF, we need to set an observation error variance (OEV) for each observed data point to quantify the uncertainty of the observation. Because only one data point is observed in each location at a given time (no repeated measurement), it is difficult to directly estimate OEV. Here, we assumed a heuristic form of OEV: $\sigma_{lt}^{2} = \sigma_{l0}^{2} + (y_{lt}^{o})^{2}/h$, where $\sigma_{l0}^{2}$ is the baseline OEV for location $l$ and $y_{lt}^{o}$ is the observed disease data in location $l$ at time $t$. The parameter $h$ controls how the OEV scales with the magnitude of observed disease data. Similar forms of OEV have been successfully used for inference and forecasting for a range of infectious diseases (11, 34–43).

To generate probabilistic forecasts at time $t$, we initialized the ensemble of model states by randomly drawing model parameters and variables from their prior distributions. The model states were updated using the EAKF at each time step when new observations became available, until the most recent observations at the forecast time $t$. During each EAKF update, we looped through observations in all locations. The observation in location $l$ was used to update model variables and parameters within the same location and global parameters (e.g., transmission rate in the simulated outbreak). The updated ensemble of models at time $t$ was integrated into the future to generate probabilistic forecasts. In implementation, we used 100 ensemble members. Details on model initialization and parameter configuration for different viruses are provided in the following sections.

**Pseudo-code of the inference algorithm.** The inference algorithm proceeds as follows.

Determine the onset time of each location. Run the bi-linear regression model to identify the onset times in all locations and rank locations according to the chronological order of estimated onset times: $\ell_1, \ell_2, \cdots, \ell_n$.

Starting from an empty set of directed transmission links $\mathcal{P}$, we sequentially add transmission links to $\mathcal{P}$. Define a mobility network $\mathcal{G}$ that comprises the bi-directional mobility links (both directions) corresponding to all transmission links in $\mathcal{P}$. Select a set of seeding locations $\mathcal{S}$. Define $\mathcal{L}_1$ as the set of seeding locations and those that already have estimated infection sources. We initialize $\mathcal{L}_1$ using the seeding locations $\mathcal{S}$. The rest of locations that do not have estimated infection sources yet are grouped in a set $\mathcal{L}_0$. Our goal is to gradually identify the infection sources of locations in $\mathcal{L}_0$, move them into $\mathcal{L}_1$, and add the inferred transmission links to $\mathcal{P}$.

While $\mathcal{L}_0$ is not empty

Select the location $\ell \in \mathcal{L}_0$ with the earliest onset time at $t_\ell$.

For $\ell' \in \mathcal{L}_1$ *(loop through all potential infection sources, parallelled in computing)*

Define the current mobility network as $\mathcal{G}' = \mathcal{G} \cup \{\ell' \leftrightarrow \ell\}$, where $\ell' \leftrightarrow \ell$ is the mobility (both flight and commuting) between $\ell'$ and $\ell$.

Generate one-week ahead forecasts using the transmission model with mobility network $\mathcal{G}'$ at the onset time $t_\ell$.

Compute the mean forecast error for the number of infections at time $t_\ell + 1$.

Use the Mann-Whitney U test to determine if this candidate model significantly outperforms the null model (no mobility linked to $\ell$) in forecasting, using a p-value threshold of 0.05.

End For

If there exist candidate models outperforming the null model

Select the mobility network with the lowest mean forecast error $\mathcal{G}'_{best} = \mathcal{G} \cup \{\ell'_{best} \leftrightarrow \ell\}$.

Update the current set of transmission links $\mathcal{P} = \mathcal{P} \cup \{\ell'_{best} \to \ell\}$.

Update the current mobility network $\mathcal{G} = \mathcal{G} \cup \{\ell'_{best} \leftrightarrow \ell\}$.

Move location $\ell$ from $\mathcal{L}_0$ to $\mathcal{L}_1$. *(infection source was found)*

Else *(no infection source was found this round)*

Update the estimated onset time of $\ell$ as $t_\ell + 1$ (day or week).

If the onset time of location $\ell$ has been updated for $d$ times

Move location $\ell$ from $\mathcal{L}_0$ to $\mathcal{L}_1$, define location $\ell$ as a new seeding location. *(no reliable infection source was found after $d$ attempts)*

Else

Update the order of location $\ell$ in $\mathcal{L}_0$ based on the updated onset time. *(location $\ell$ will be examined again in following rounds)*

End If

End If

End While

**Remarks.** The inference framework has a few designs to address uncertainties in onset time estimation and the setting of seeding locations. First, if the inference algorithm cannot identify an infection source of a location at the estimated onset time (i.e., none of the candidate models outperformed the null model), we will update the estimated onset time and search for its infection source again in later rounds. Second, if the inference algorithm cannot identify the infection source of a location after several attempts (8 attempts for the daily COVID-19 data, 4 attempts for the weekly influenza data), we treat this location as a location without reliably identified infection sources. The major computation burden is the large number of forecasts using candidate models.

To speed-up the computation, we parallelled the forecasting using candidate models as they can be performed independently.

**Ensemble inference.** As the transmission model has stochastic dynamics, different realizations of the inference algorithm may result in different transmission networks. Performing independent realizations of inference provide a means of quantifying the uncertainty of the inferred transmission links. In the following analyses, we performed 100 independent realizations of inference and formed an ensemble of reconstructed transmission networks. For each transmission link, we recorded its occurrence probability in the ensemble of inferred transmission networks (i.e., the percentage of networks including that specific transmission link). We found that a transmission link that occurred more frequently in the ensemble had a higher inference accuracy. To select high-confidence transmission links, we used a threshold for the occurrence probability in the ensemble to reconstruct the transmission network. Transmission links with occurrence probabilities higher than the threshold were included in the final transmission network. The threshold was optimized using F1-score to balance the precision and recall in the inference results (see details in the next section).

**Validation using simulated outbreaks**

We validated the inference framework using simulated outbreaks, where the ground-truth transmission links were known. Specifically, we used the simulated outbreak originated from the Minneapolis–St. Paul–Bloomington, MN–WI MSA, as shown in Fig. 1A. Weekly new infections in all MSAs generated from this specific simulation realization were used in the inference. By running bi-linear regression, we identified that the Minneapolis–St. Paul–Bloomington, MN–WI MSA had the earliest onset time. This MSA was set as the seeding location in the inference. To initialize the ensemble of model states in the inference system, we randomly drew initial infections in the seeding location from a uniform distribution $U[10,20]$. Other parameters were drawn from their prior distributions $\beta \in U[0.3,0.45]$, $D \in U[3,5]$ days, and $L \in U[1,2]$ years. The dispersion parameter was set as $r = 2.36$. In the EAKF, the OEV for the observation from location $l$ and day $t$ was set as $OEV_{lt} = 10{,}000 + (y_{lt}^o)^2/4$, where $y_{lt}^o$ is the number of new infections in location $l$ and day $t$. In the inference, each location had up to 4 attempts to infer infection source ($d = 4$ weeks).

Since the inference algorithm includes stochastic components, outcomes of independent realizations of inference may be different. To capture this uncertainty, we performed 100 independent realizations of the inference. For the reconstructed transmission network in each inference, we computed the precision (the percentage of correctly identified positives out of all predicted positives) and recall (the percentage of correctly identified positives out of all actual positives) using the ground-truth transmission links in Fig. 1A. Here, an inferred transmission link was accurate if both the infection source and mobility mode were correctly identified. The distributions of the precision and recall for single inference realizations are shown in Fig. 2A.

The 100 realizations of inference produced an ensemble of transmission networks. For the transmission links appearing in these networks, we used a threshold for their occurrence probabilities to select a high-confidence transmission network. To find the optimal threshold, we tested all possible values (from 0% to 100%) and computed the F1-score, the harmonic mean of the precision and recall of the resulted transmission network. The threshold achieving the maximum F1-score was selected to balance the precision and recall.

**Robustness of the inference framework**

**Robustness to the selection of seeding locations.** We first examined the robustness of findings to the selection of seeding locations. In the inference, we additionally included New York-Newark-Jersey City, NY-NJ-PA MSA, another location with early onset time, in the seeding locations. While the selected seeding locations were not correct, the precision and recall for each individual inference remained similar (Fig. S8A). The performance of the ensemble inference did not change substantially, producing a slightly higher optimal threshold (i.e., the inference was more conservative) (Fig. S8C).

**Robustness to model misspecification.** In the previous inference, the transmission model used in the inference was the same as the one generating the outbreak data. In reality, however, the transmission model is mis-specified compared to the actual epidemic process. To examine whether the inference can yield satisfactory results when the model cannot fully describe the epidemic process, we performed the following analysis. We randomly removed 10% of cross-MSA mobility (both commuter and airline travel were removed if a pair of MSAs was selected) from the transmission model and generated outbreak data as the ground truth. In the inference, we still used the transmission model with full mobility, different from the model that generated disease data. The precision and recall for each individual inference dropped modestly (Fig. S8B). However, the precision and recall in the ensemble inference remained above 80% (Fig. S8D), suggesting a robust performance with model misspecification.

**Robustness to data missingness.** For H1N1pdm influenza, ILI+ incidence data were only available in 220 MSAs. As data in other MSAs were missing, we used a transmission model that only included 220 MSAs in the inference. To examine how this choice would affect the inference outcome, we performed another sensitivity analysis. We simulated an outbreak using the full transmission model with 367 MSAs and configurations for H1N1pdm influenza. Then we performed inference using a transmission model with only 220 MSAs and the disease data from these locations. We computed the precision and recall using the ground-truth transmission links between the included 220 MSAs. For each individual inference, the precision and recall were 65.3% (95% CI [57.5%, 71.1%]) and 63.4% (95% CI [56.1%, 69.0%]), respectively (Fig. S9B). The ensemble inference achieved a precision of 81% and a recall of 77% (Fig. S9D). This analysis indicates that the inference using disease data from a subset of locations still had acceptable performance.

**Remarks.** For each location, the inference framework aims to identify the most important mobility link from potential infection sources that can support the first-order approximation of the infection in the focal location. The goal in each round of inference is not to accurately forecast the infection, but to find the mobility link leading to the lowest forecast error (i.e., the candidate model does not necessarily accurately predict the observation. It can generate biased forecasts but needs to outperform other candidate models). This relatively flexible criteria may allow the inference to tolerate errors in the transmission model construct and initial settings. In addition, the EAKF can dynamically adjust epidemiological parameters (e.g., the transmission rate) and state variables (e.g., susceptible and infected populations) based on most recent observations, partly mitigating the errors caused by model misspecification.

**Inference for SARS-CoV-2**

We employed the inference system to infer the spatial spread of SARS-CoV-2 in the US. Due to limited testing capacity and the existence of asymptomatic and mild infections, there were severe underreporting of COVID-19 infections. To address this issue, instead of using the daily confirmed COVID-19 cases, we used the estimated daily new COVID-19 infections (both reported and unreported) in 3,142 US counties in Ref. (1). We aggregated the county-level daily infection data to the MSA scale.

In the inference, we used a transmission model with an SEIRS (susceptible-exposed-infectious-recovered-susceptible) dynamics.

$$\frac{dS_i}{dt} = -S_i \sum_j \frac{\beta_j M_{j\leftarrow i} I_j}{N_j} + \frac{R_i}{L} - \frac{S_i}{N_i} \sum_{j\neq i} C_{j\leftarrow i} + \sum_{j\neq i} \frac{C_{i\leftarrow j} S_j}{N_j}, \quad [16]$$

$$\frac{dE_i}{dt} = S_i \sum_j \frac{\beta_j M_{j\leftarrow i} I_j}{N_j} - \frac{E_i}{Z} - \frac{E_i}{N_i} \sum_{j\neq i} C_{j\leftarrow i} + \sum_{j\neq i} \frac{C_{i\leftarrow j} E_j}{N_j}, \quad [17]$$

$$\frac{dI_i}{dt} = \frac{E_i}{Z} - \frac{I_i}{D} - \frac{I_i}{N_i}\sum_{j\neq i} C_{j\leftarrow i} + \sum_{j\neq i} \frac{C_{i\leftarrow j} I_j}{N_j}, \quad [18]$$

$$\frac{dN_i}{dt} = -\sum_{j\neq i} C_{j\leftarrow i} + \sum_{j\neq i} C_{i\leftarrow j}. \quad [19]$$

Here $E_i$ is the exposed population in location $i$ and $Z$ is the mean latency period. Other parameters are the same as in Eqs. [2-4]. Disease-related epidemiological parameters in each EAKF ensemble member were randomly drawn from the following distributions and fixed during the model calibration: $Z \in U[3,4]$ days (44), $D \in U[3,5]$ days (45), and $L \in [2,10]$ years. A relatively long duration of immunity was used as few reinfections occurred during the early phase of the COVID-19 pandemic. The dispersion parameter for SARS-CoV-2 was set as $r = 0.55$ (22). To represent the heterogeneity of disease transmission across locations, we defined location-specific transmission rates $\beta_i$ and allowed them to change over time (updated on each day using the EAKF), following Ref. (1). The prior distribution of the transmission rates was set as a uniform distribution $\beta_i \in U[1.3,1.7]$. In the EAKF, the OEV for the observation from location $l$ and day $t$ was set as $OEV_{lt} = 100 + (y_{lt}^o)^2/100$, where $y_{lt}^o$ is the number of new infections in location $l$ and day $t$. In the inference, each location had up to 8 attempts to infer infection source ($d = 8$ days).

Using the bi-linear regression method, we estimated the onset times of location transmission for all MSAs. We found that the Seattle–Tacoma–Bellevue, WA MSA and the New York-Newark-Jersey City, NY-NJ-PA MSA had the earliest onset time and set both MSAs as the seeding locations in the inference. This choice of seeding locations is consistent with findings using SARS-CoV-2 genomic data (46). We performed inference from February 21st, 2020 to May 10th, 2020. In this model, we defined the number of daily new infections as the number of individuals transitioning from susceptible (S) to exposed (E) status. To initialize the model on the first day of inference, for each ensemble member in the EAKF, we randomly drew the total number of infectious individuals from $U[500,1000]$ and distributed them to the two seeding locations according to the cumulative numbers of infections until March 5th in the two locations (estimated 12,774 infections in Seattle–Tacoma–Bellevue, WA MSA and 16,331 infections in New York-Newark-Jersey City, NY-NJ-PA MSA). The range of initial infections were roughly estimated using the number of daily new infections in the two MSAs. At the beginning of the inference, the two MSAs had a total of ~200 estimated daily new infections. Considering the duration of the infectious period of 3 to 5 days, using a range of 500 to 1000 total infectious individuals is reasonable. As infectious individuals were people who had transitioned from the exposed state, we set the exposed population in each ensemble member the same as the infectious population. The rest of population was set as susceptible. Note the EAKF can dynamically adjust model variables based on observations, partially mitigating the errors in the initial seeding. This crude calculation has been used to initialize SARS-CoV-2 models in previous studies (1, 11, 45).

To identify the optimal threshold for occurrence probabilities in the ensemble inference, we simulated an outbreak using the configurations for SARS-CoV-2 and performed 100 inferences (Fig. S9A). For this example, the optimal threshold was 46% (Fig. S9C), which we used in the inference using real-world SARS-CoV-2 data. We also presented the transmission networks inferred using three other thresholds: 40%, 60%, and 80% (Fig. S14).

As a sensitivity analysis, we performed inference using another set of seeding locations: Seattle–Tacoma–Bellevue, WA MSA, New York-Newark-Jersey City, NY-NJ-PA MSA, and San Francisco-Oakland-Hayward, CA MSA. The reconstructed transmission network remained similar (Fig. S15).

Reinfection is an important issue over longer time scale. Indeed, immune waning can be both continuous and impulsive, and previously infected individuals might be less susceptible to reinfection, and reinfection might be less infectious in general. However, in this study, we only simulated the initial stage of the COVID-19 pandemic; therefore, the impact of immunity duration should be limited. We did test a shorter immunity duration $L = 0.5$ year, and the reconstructed network remained similar (Fig. S16).

**Inference for H1N1pdm influenza**

We used the transmission model in Eqs. [2-4] for 220 MSAs to infer the transmission network of H1N1pdm influenza. In the inference, we need to map the observed ILI+ incidence indicator to the weekly new infections in the model. Denote $p(i|m)$ as the probability a person seeking medical treatment, $m$, has ILI symptoms (we leave out the indices of location and time for notational convenience). That is $p(i|m) = visit_{ILI}/visit_{All}$. Denote the number of influenza infections in an MSA as

$$flu = p(i) \times pop \times pr, \quad [20]$$

where $p(i)$ is the probability of an individual having ILI symptoms, $pop$ is the population size of the MSA, and $pr$ is the concurrent positivity rate. By Bayes' rule, $p(i|m) = p(i)p(m|i)/p(m)$, where $p(m|i)$ is the probability of seeking medical attention given ILI symptoms and $p(m)$ is the probability that anyone seeks medical attention for any reason (47). Recall that

$$ILI +_{incidence} = \frac{visit_{ILI}}{visit_{All}} \times \frac{pop}{100{,}000} \times pr = p(i|m) \times \frac{pop}{100{,}000} \times pr. \quad [21]$$

We have

$$flu = \gamma ILI+_{incidence} \times 100{,}000, \quad [22]$$

where $\gamma = p(m)/p(m|i)$. The parameter $\gamma$ is typically not observable. To estimate this parameter, we used the serological survey for H1N1pdm influenza (48). By December 2020, the estimated incidence of natural infection accounted for 20.2% (95% CI [10.1%, 28.3%]) of US population. We estimated the parameter $\gamma \approx \sum flu / (\sum ILI+_{incidence} \times 100{,}000)$, where $\sum flu$ is the total number of infections estimated from the serological study and $\sum ILI+_{incidence}$ is the cumulative ILI+ incidence by December 2009. The range of $\gamma$ is from 0.115 to 0.323. In the inference, we set $\gamma = 0.2$ and converted the $ILI+_{incidence}$ data to the weekly number of infections, which we used as the observation in the inference algorithm. Disease data from March 22nd, 2009 to July 4th, 2009 were used in the inference.

In the transmission model, parameters in each EAKF ensemble member were randomly drawn from the following distributions: $D \in U[1.9, 6]$ days (49), and $L \in U[2, 10]$ years. The choice of immunity duration should not substantially affect the model dynamics as the simulation period was much shorter than the temporal scale of $L$. The dispersion parameter for H1N1pdm influenza was set as $r = 2.36$ (26). We defined location-specific transmission rates and allowed them to change over time. The prior distribution of the transmission rates was set as a uniform distribution $U[0.2, 0.6]$. In the EAKF, the OEV for the observation from location $l$ and week $t$ was set as $OEV_{lt} = 25 + (y_{lt}^{o})^2/4$, where $y_{lt}^{o}$ is the number of new infections in location $l$ and week $t$. In the inference, each location had up to 4 attempts to infer infection source ($d = 4$ weeks).

To select seeding locations, we estimated the onset time for each MSA. As the ILI+ incidence was aggregated weekly, large uncertainty may exist due to data sparsity. Here, we selected seeding locations combining the early case reporting data from the CDC (50) and the estimated onset time. Sensitivity analysis was performed on the seeding location selection. Three MSAs with early reported cases were set as the seeding locations: New York-Northern New Jersey-Long Island, NY-NJ-PA MSA, San Antonio, TX MSA, San Diego-Carlsbad-San Marcos, CA MSA. The estimated onset times for New York-Northern New Jersey-Long Island, NY-NJ-PA MSA and San Antonio, TX MSA were ranked in the top two among 220 MSAs. We randomly drew initial infections from the distribution $U[1500, 3000]$ and distributed them to the three locations based on the reported cases as of April 28th, 2009 (California (10), New York (45), and Texas (6)). The range of initial infections was roughly estimated as follows. In the week of April 19th, about 50 cases has been confirmed (50, 51). Assuming a 1% reporting rate and a two-week reporting delay, ~ 5000 infections occurred in the week of April 5th. Using a doubling time of one week (52), the number of infections in the week of March 22nd was ~1100. Considering an infectious period up to 6 days, setting the initial infected population on March 22nd at the order of $O(10^3)$ is reasonable. We estimated the optimal threshold in the ensemble inference using a simulated outbreak with configurations for H1N1pdm influenza (Fig. S9D). We also presented the transmission networks inferred using three other thresholds: 40%, 60%, and 80% (Fig. S14). As a sensitivity analysis, we performed inference by adding another three locations with early estimated onset times to the initial sites: Terre Haute, IN MSA, Fort Wayne, IN MSA, and Springfield, IL MSA. The reconstructed transmission network remained similar (Fig. S15). We also performed another inference using a different distribution of initial infections $U[100,1000]$ and found the reconstructed transmission network was similar (Fig. S17).

To assess the effect of increasing detection in early weeks, we performed sensitivity analyses assuming a weekly 20% or 40% increase in the detection rate during the first five weeks of the pandemic, before reaching a plateau in week six. Specifically, we modified the ILI+ time series to adjust for this under-detection (i.e., the ILI+ in each MSA in week $t$ was multiplied by $1/(80\%)^{6-t}$ or $1/(60\%)^{6-t}$ for $1 \leq t \leq 5$) and re-ran the inference. The resulting transmission networks remained structurally similar except a few early, long-range transmission pathways (Fig. S18). These altered transmission links were originally uncertain with occurrence probabilities close to the selection threshold. For instance, the infection source of Los Angeles switched from San Antonio to New York, where the new occurrence probability for the link from New York to Los Angeles was 45%, close to the threshold of 42%. The major infection source of San Francisco switched from San Diego to New York, where, in the new inference, the occurrence probabilities for the links from New York and San Diego were 49% and 46%, both above the threshold of 42%. (Note, in the figure we only visualized the transmission link from the inferred major infection source.) For Seattle and Portland, their infection sources switched from New York to San Diego.

**Simulations of future pandemics**

For a future respiratory pandemic with unknown characteristics, we performed simulations under a variety of scenarios on outbreak origins, virus transmissibility, and superspreading potentials. Specifically, we considered outbreaks originating within the US and those introduced into the US. For outbreak origins within the US, we selected 10 MSAs with large numbers of reported H5N1 HPAI spillovers (27). Origins for pathogens introduced from other regions were selected as major airline transportation hubs. The list of selected outbreak origins is provided in Table S2. In the simulation, we assume each outbreak has one origin. The transmission model follows a susceptible-infected-recovered dynamics described in Eqs. [2-4]. To represent various virus transmissibility, we used 7 values for the transmission rate $\beta$, ranging from 0.4 to 1 with a step of 0.1. Using an average infectious period of 4 days, this range translates to a basic reproductive number between 1.6 and 4. For superspreading potentials, we used 7 different dispersion parameters $r$ from 0.01 to 10, evenly distributed on the log scale. In total, we simulated 980 different scenarios with the combination of outbreak origin, virus transmissibility, and superspreading potential.

**Estimating infection source using mobility heuristics**

We used two heuristic approaches to identify the infection source of each MSA: 1) the volume of incoming mobility, and 2) the expected number of imported infections.

**The volume of incoming mobility.** For each mobility mode (commuting or flight), denote $F_{i\leftarrow j}^{t}$ as the number of individuals traveling from location $j$ to $i$ at time $t$. The cumulative volume of incoming mobility from location $j$ to location $i$ until the onset time of $i$ is defined as $\sum_{t=1}^{t_{onset}} F_{i\leftarrow j}^{t}$, where $t_{onset}$ is the onset time of location $i$. Based on this definition, the infection origin of MSA $i$ is estimated to be the MSA contributing to the largest cumulative mobility volume to $i$ (combining commuting and flight). The transportation mode for each transmission link is defined as the one contributing to the majority of mobility volume.

**The expected number of imported infections.** For each MSA $i$, we define the infection importation risk using the number of expected cumulative infections introduced through commuting or airline travel from other locations. The infection importation risk from location $j$ to location $i$ through a certain mobility mode was estimated by

$$\hat{F}_{i\leftarrow j} = \sum_{t=1}^{t_{onset}} \sum_{j=1}^{n} F_{i\leftarrow j}^{t} \times \frac{y_{jt}^{o}}{N_j}, \quad [23]$$

where $y_{jt}^{o}$ is the number of new infections in location $j$ at time $t$ (daily for SARS-CoV-2, weekly for H1N1pdm influenza), $N_j$ is the population in location $j$, $n$ is the total number of locations, and $t_{onset}$ is the onset time of location $i$. To estimate the relative infection importation risk from other locations to $i$, we normalized $\hat{F}_{i\leftarrow j}$ across all potential infection sources and defined the infection importation risk used in Fig. 4 as

$$f_{i\leftarrow j} = \frac{\hat{F}_{i\leftarrow j}}{\sum_k \hat{F}_{i\leftarrow k}}. \quad [24]$$

Note the infection importation risk was defined for each MSA to estimate the most likely infection source given mobility flows and infection status in other locations.

**Modeling surveillance and intervention strategies for future pandemics**

We performed simulations to assess the effects of wastewater surveillance among air travelers and subsequent interventions to slow down the spatial spread of a novel pathogen. Since the cryptic transmission in the outbreak origin is difficult to detect, we deployed wastewater surveillance in transmission hubs identified in Fig. 4B, agnostic of the origin of the novel pathogen. Specifically, we selected a given number of transmission hubs following a decreasing order of the probabilities of each MSA infecting ≥ 5 other MSAs among the simulations in Fig. 4B. Previous research estimated that the detection rate of an infected individual through wastewater surveillance on flights is in the range of 11-22% for SARS-CoV-2 (53). In our simulations, we used a 20% detection rate for each infected individual among air travelers arriving at the MSAs with wastewater surveillance. Interventions were enacted when at least one infection was detected using wastewater surveillance, or the cumulative local infections in an MSA reached a threshold (e.g., some locally infected people may seek medical treatment and be discovered at hospitals), whichever came first. We assume the interventions will reduce the local transmission rate by a certain percentage due to risk aversion, social distancing, or other control policies. Here we assume a relatively mild disease severity and set the threshold of community transmission detection as 5,000 cumulative infections in an MSA. We also tested a threshold of 1,000 cumulative infections as a sensitivity analysis (Fig. S20).

We simulated a hypothetical outbreak originating from New York, using model parameters and initial seeding number same as in Fig. 1A ($R_0 = 1.5$, $r = 2.36$), and examined the number of MSAs with local onsets in the first 60 days, varying the number of wastewater surveillance sites and the percentage reduction of local transmission rates when interventions are enacted. In Fig. 4G, we tested 0, 1, 5, 10 and 20 surveillance sites and 5%, 10%, 15%, 20% and 25% reduction of transmission rates. Due to stochastic dynamics, the outcomes of independent realizations may be different. For each strategy, we repeated the simulation for 100 times and recorded the number of MSAs with local onsets by the day 60. The distributions of outcomes are shown in Fig. S19. In Fig. 4H, we fixed the transmission rate reduction at 25% and tested the number of surveillance sites from 0 to 30. In Fig. S20, we repeated the analysis but used a threshold of 1,000 cumulative infections to detect community transmission. We also tested another outbreak originating from Minnesota, with configurations same to Fig. 1A, yielding similar results (Fig. S21).

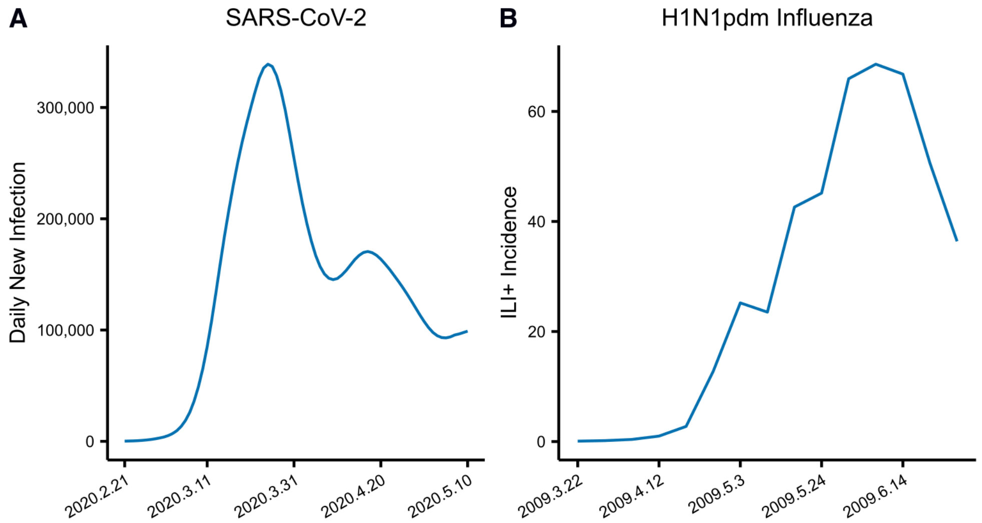


**Fig. S1. National time series for SARS-CoV-2 and H1N1pdm influenza**. (**A**). The estimated daily new infection numbers in 375 MSAs in the US. (**B**). The weekly ILI+ incidence indicator in 220 MSAs in the US.

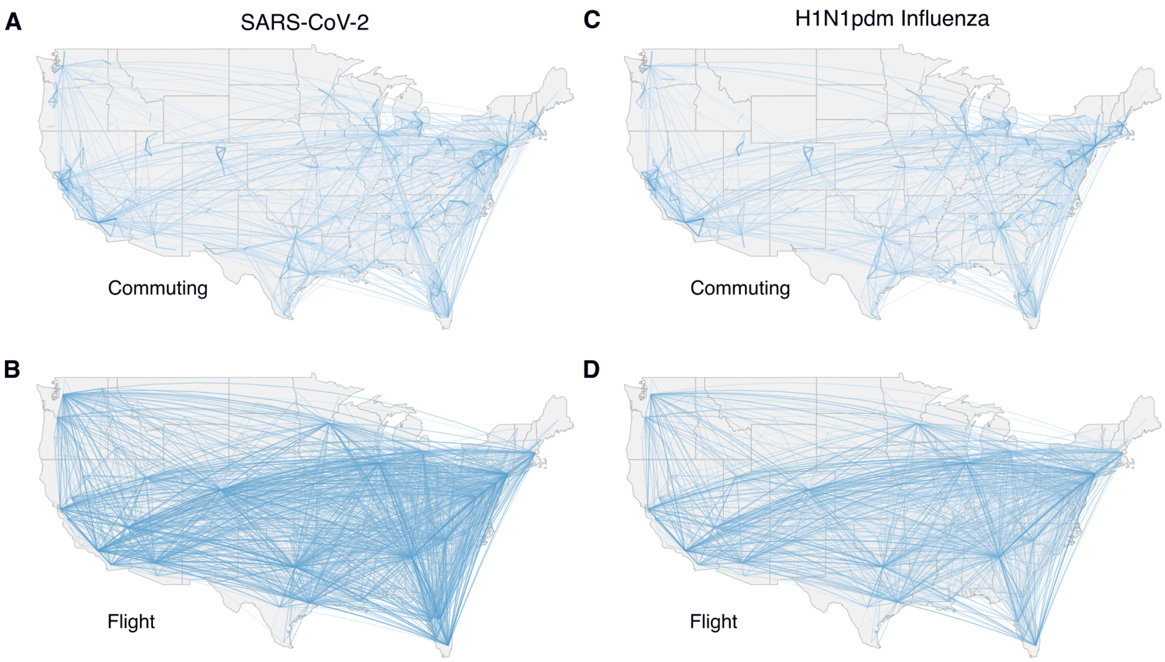


**Fig. S2. Human mobility during the COVID-19 and 2009 H1N1 influenza pandemics**. (**A**). Daily commuting between MSAs from the 2016-2020 5-Year ACS Commuting Flows. The width of links represents the mean volume of commuting averaged over both directions. (**B**). Average daily volume of inter-MSA airline travel in 2020 from the US Bureau of Transportation Statistics. The daily number of passengers between MSAs is averaged over all days in 2020 and both directions of airline travel. (**C**). Daily commuting between MSAs from the 2009-2013 5-Year American Community Survey. (**D**). Average daily volume of inter-MSA airline travel in 2009 from the US Bureau of Transportation Statistics.

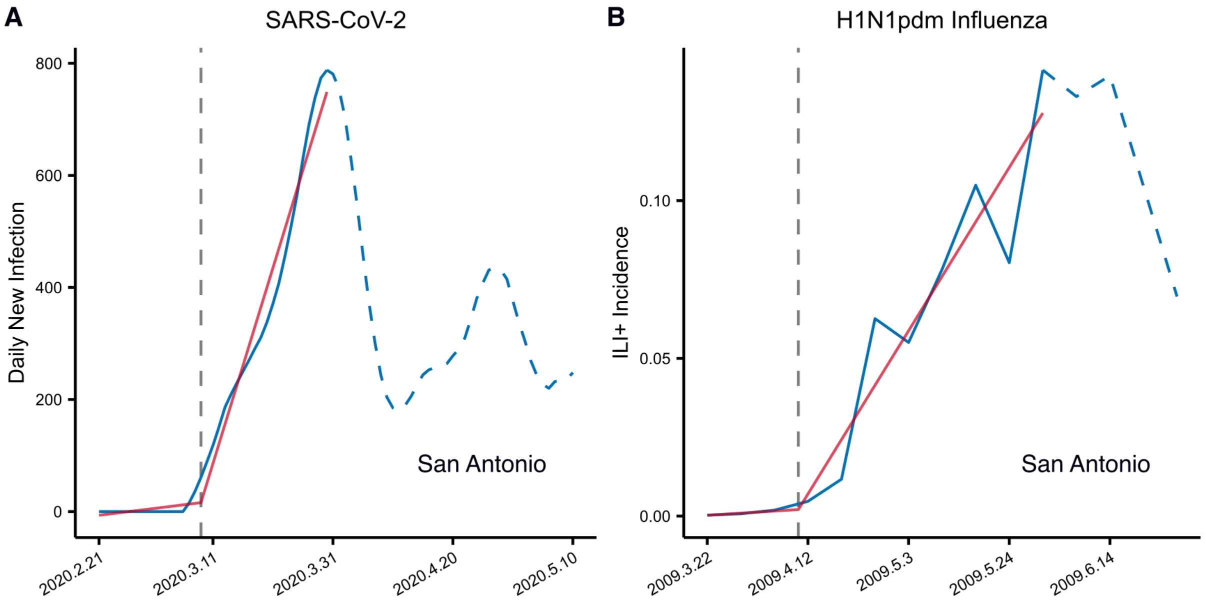


**Fig. S3. Estimation of onset time using bi-linear regression.** We used a bi-linear regression to identify the break point of the disease time series. (**A**). An example for SARS-CoV-2 in San Antonio–New Braunfels, TX MSA. The solid blue line shows the data used in the bi-linear regression. The red line is the fitted line. The vertical dash line indicates the identified onset time. (**B**). An example for H1N1pdm influenza in San Antonio. Weekly ILI+ incidence time series was used to identify local onset time.

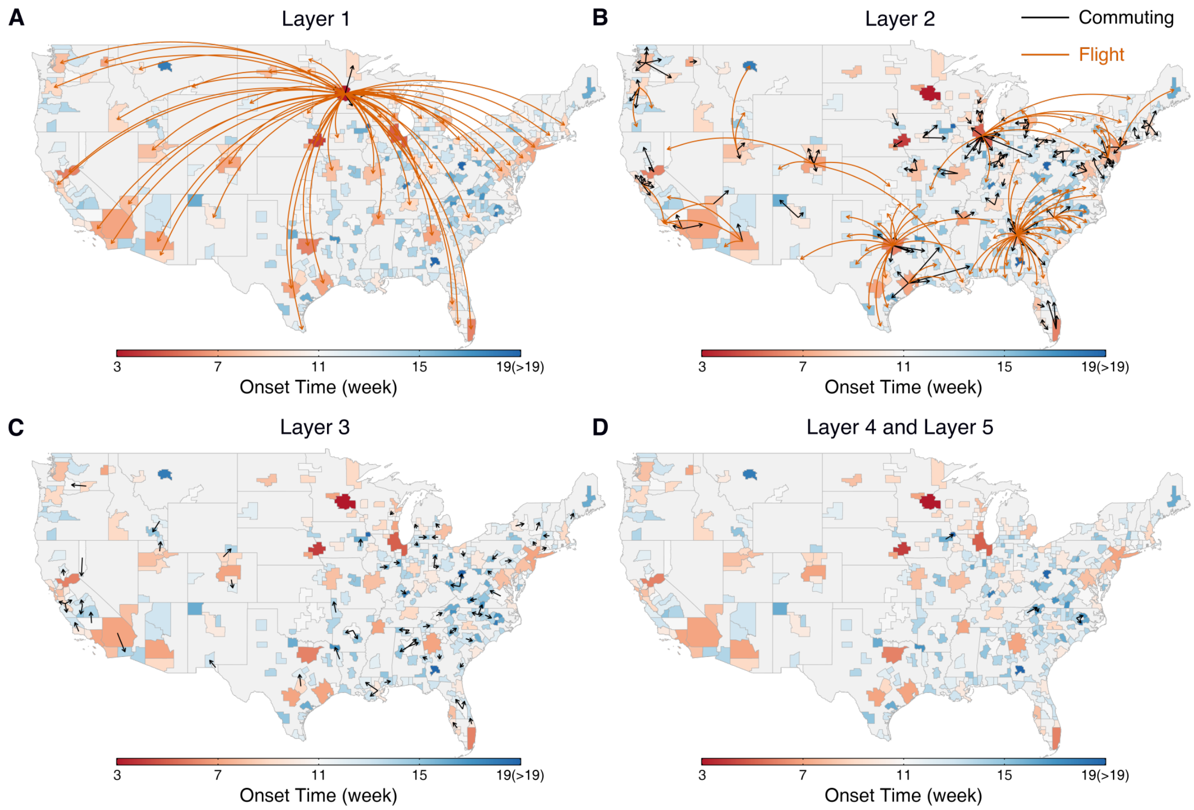


**Fig. S4. The spatial hierarchical structure of the spatial spread of a simulated outbreak.** We visualize the transmission links across MSAs at different layers from the origin MSA in Minnesota for the simulated outbreak in Fig. 1A. Most transmission through airline travel occurred in the first two layers. Transmission through commuting mostly occurred in the second and third layers.

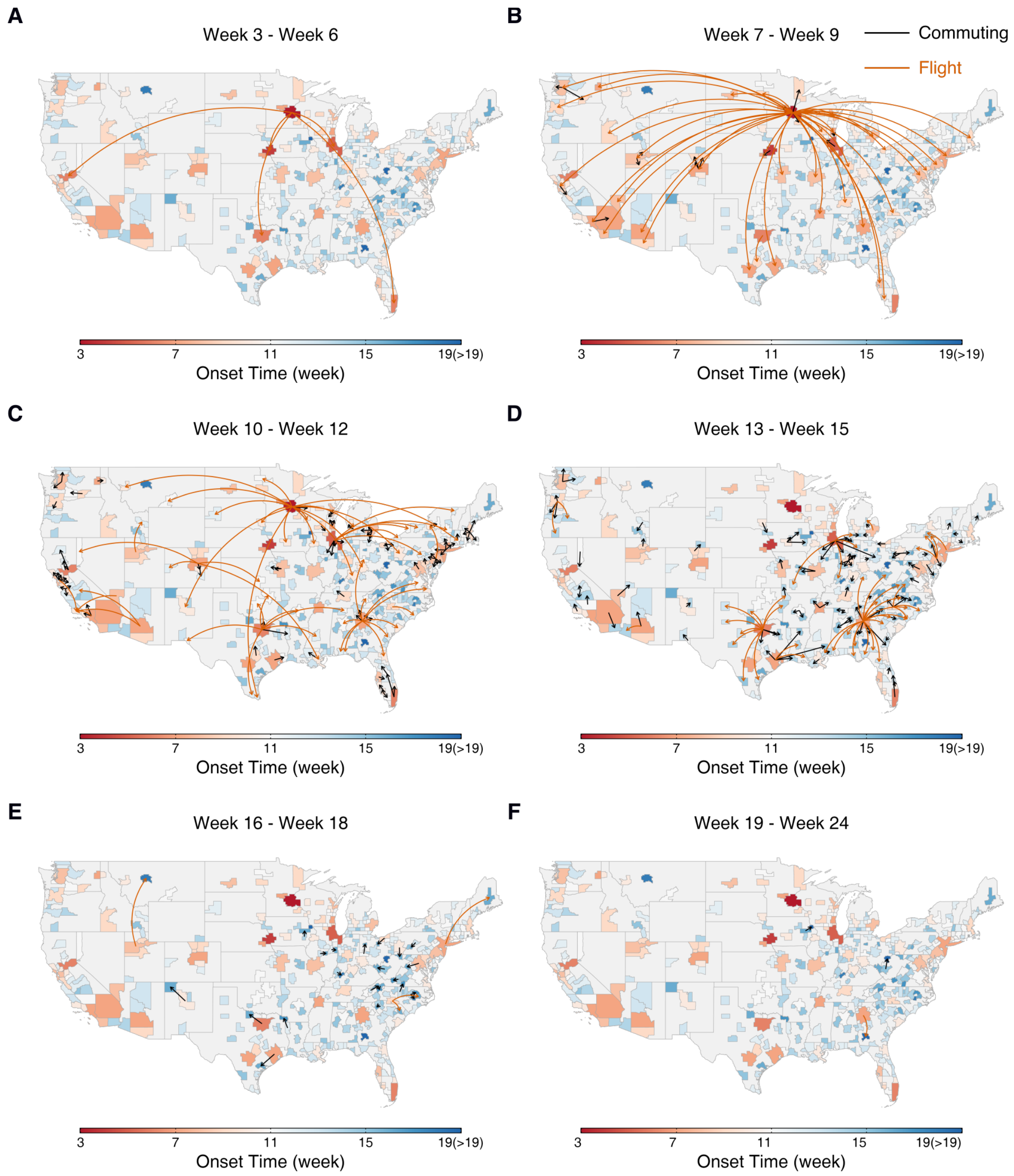


**Fig. S5. The spatial spread of a simulated outbreak over time.** We show the transmission links occurred during a given time interval for the simulated outbreak in Fig. 1A. The week numbers reflect the time since the start of the simulation.

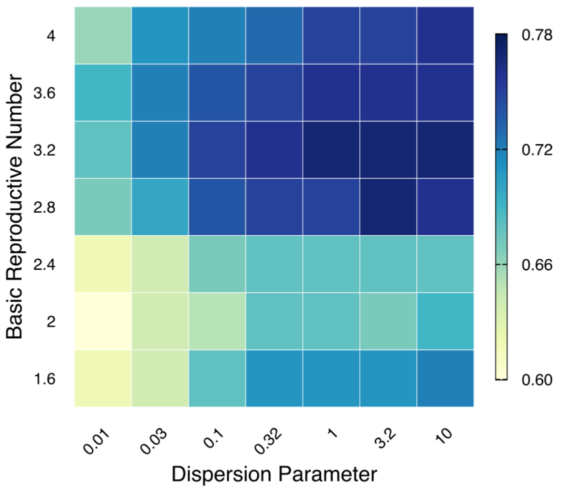


**Fig. S6. The uncertainty of transmission networks for simulated outbreaks.** We generated synthetic outbreaks using different transmission rates and dispersion parameters, with other settings same as the simulation in Fig. 1A. For each parameter combination, 100 independent simulations were performed. The color shows the fraction of stable transmission links that appeared in at least 80% of simulations.

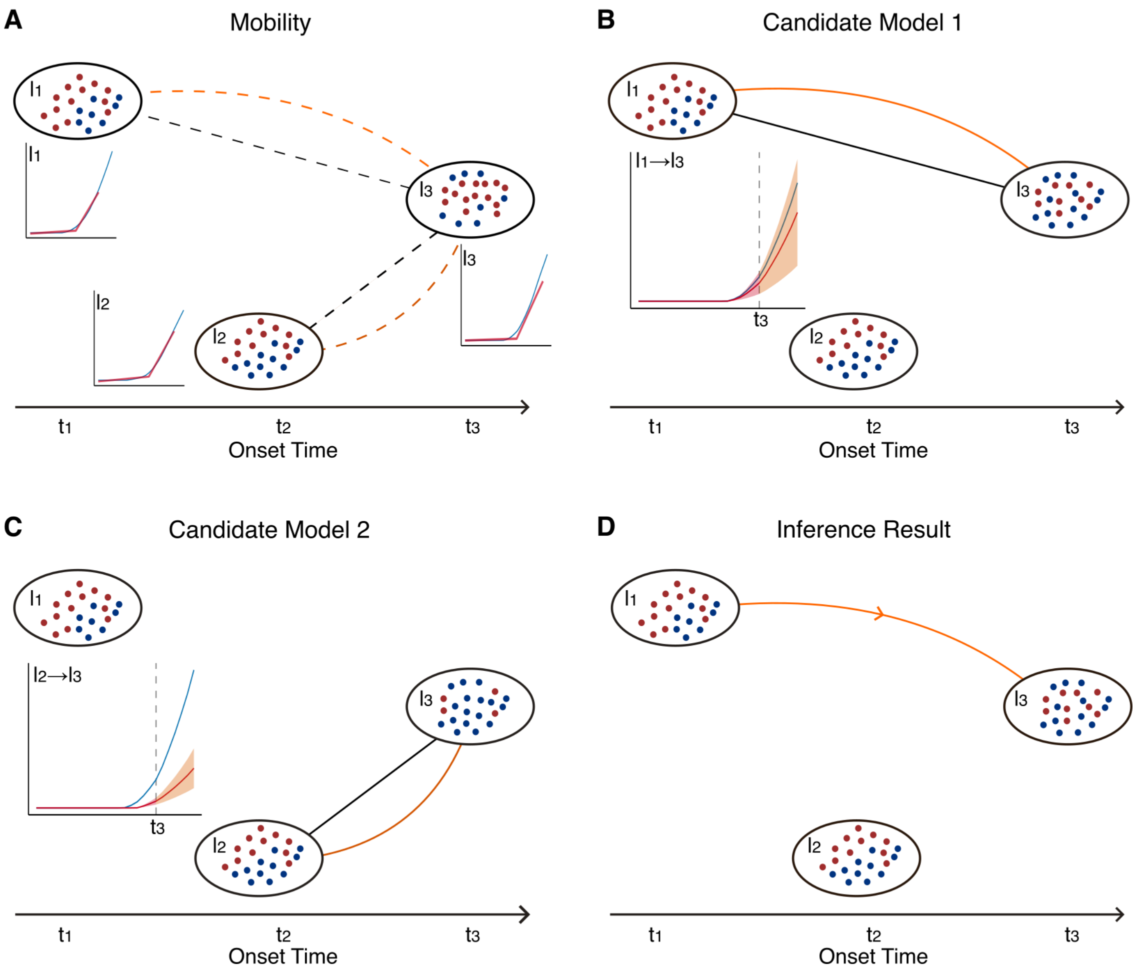


**Fig. S7. An example of the inference algorithm for three locations.** (**A**). For three locations $l_1$, $l_2$, and $l_3$, we estimated their onset times at $t_1$, $t_2$, and $t_3$ ($t_1 < t_2 < t_3$). Suppose that $l_1$ and $l_2$ are seeding locations, we aim to infer the infection source of location $l_3$. The dashed lines represent mobility links between locations, where straight lines represent commuting and curved lines represent airline travel. (**B**). Candidate model 1 with only mobility between $l_1$ and $l_3$. We calibrate candidate model 1 until the onset time of location $l_3$ and generate the forecast for the next time step. The forecast agrees well with the observation (inset). (**C**). Candidate model 2 with only mobility between $l_2$ and $l_3$. We calibrate candidate model 2 until the onset time of location $l_3$ and generate forecasts for the next time step. The forecast deviates from the observation (inset). (**D**). As the candidate model 1 better predicts the infection in location $l_3$ and significantly outperforms the null model (i.e., no mobility links to $l_3$), we infer the infection source of location $l_3$ as $l_1$. We estimate that the cumulative number of infections attributable to airline travel from $l_1$ to $l_3$ until $t_3$ is larger than that through commuting. Therefore, we create a transmission link from $l_1$ to $l_3$ via airline travel.

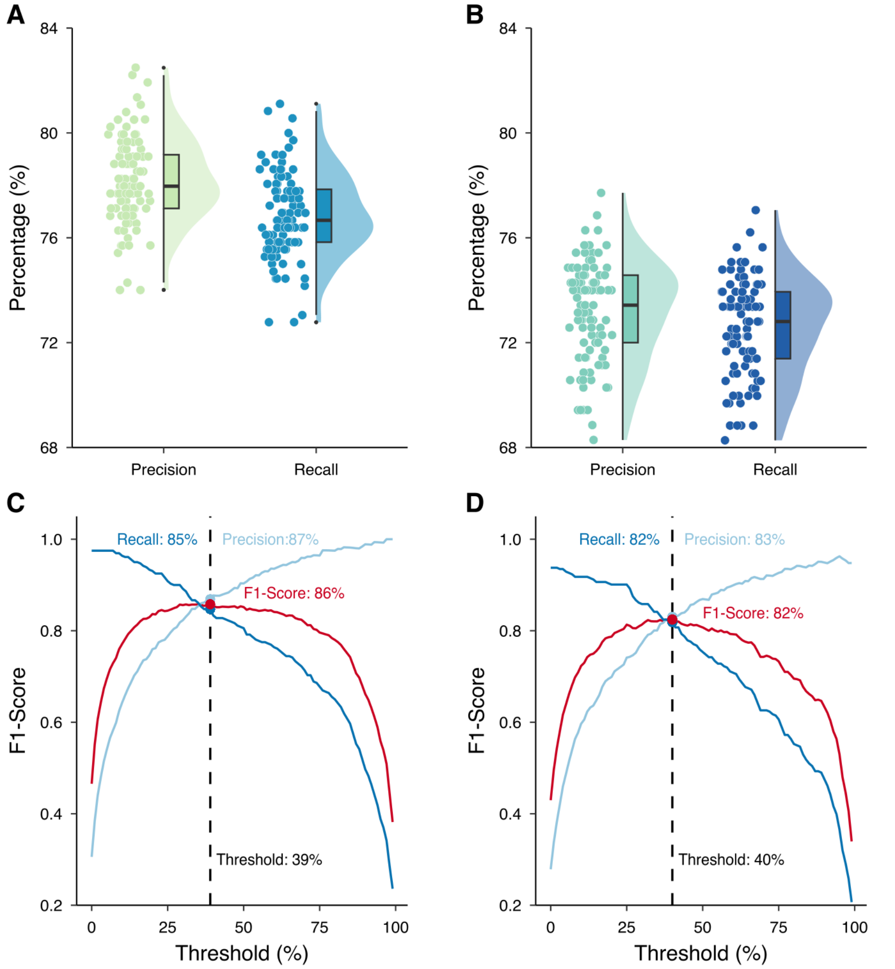


**Fig. S8. Sensitivity analyses of the inference framework for simulated outbreaks.** We performed sensitivity analyses of the inference for simulated outbreaks. In the first analysis, we used an additional location as the epidemic origin in the inference. In the second, we performed inference using a transmission model that was different from the model that generated the ground-truth outbreak data. (**A**). The precision and recall of inference results for the simulated outbreak in Fig. 1A using an additional origin MSA (New York-Newark-Jersey City, NY-NJ-PA MSA) in the inference. We performed 100 independent realizations of inference. Each dot represents the performance for one realization. Boxes show the medians and interquartile of the distributions. (**B**). The precision and recall of inference results for a simulated outbreak generated using the mobility network where 10% of mobility links were randomly removed. The inference algorithm used the full mobility network thus the transmission model in the inference was mis-specified. (**C**). The performance of the ensemble inference in (A) using different thresholds of the occurrence probability to define transmission links. (**D**). The performance of the ensemble inference in (B) using different thresholds of the occurrence probability to define transmission links.

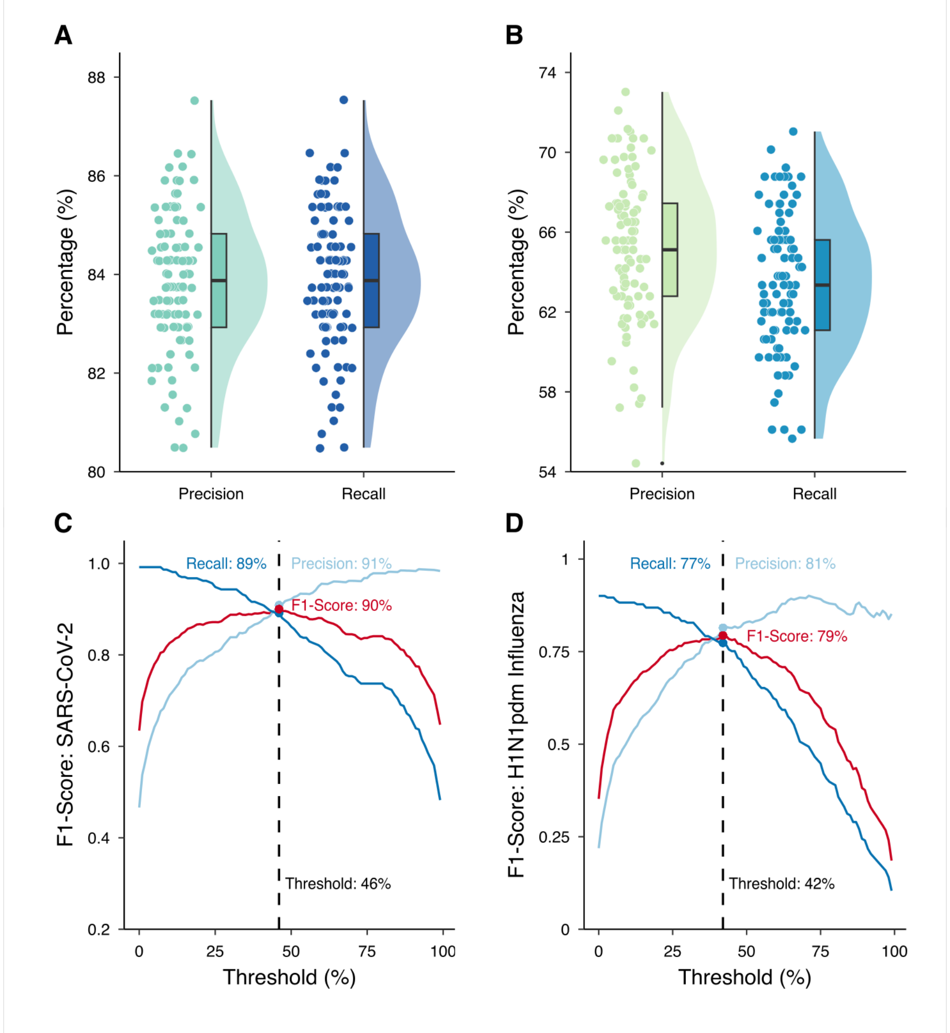


**Fig. S9. Performance of inference for simulated outbreaks tailored to SARS-CoV-2 and H1N1pdm influenza.** (**A**). The performance of inference for a simulated outbreak in all MSAs generated using parameters for SARS-CoV-2. We performed 100 independent realizations of inference. Each dot represents the performance for one realization. Boxes show the medians and interquartile of the distributions and whiskers show 95% CIs. (**B**). We generated a simulated outbreak in all 367 MSAs using parameters tailored to H1N1pdm influenza and performed inference using a subset of 220 MSAs. The precision and recall of the inference results for the subset of the 220 MSAs are presented. (**C**). The optimal threshold in the ensemble inference for the simulated outbreak using parameters for SARS-CoV-2. (**D**). The optimal threshold in the ensemble inference for the simulated outbreak tailored to H1N1pdm influenza.

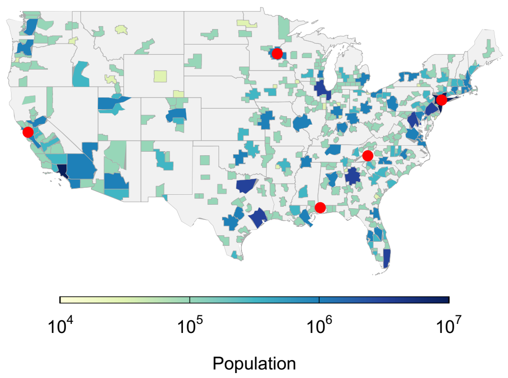


**Fig. S10. Simulation of an outbreak with multiple introductions.** Red dots on the map show the geographical distribution of the five outbreak origins used in the model simulation: 1) Mobile, AL MSA; 2) Asheville, NC MSA; 3) San Francisco-Oakland-Fremont, CA MSA; 4) Minneapolis-St. Paul-Bloomington, MN-WI MSA; 5) New York-Northern New Jersey-Long Island, NY-NJ-PA MSA.

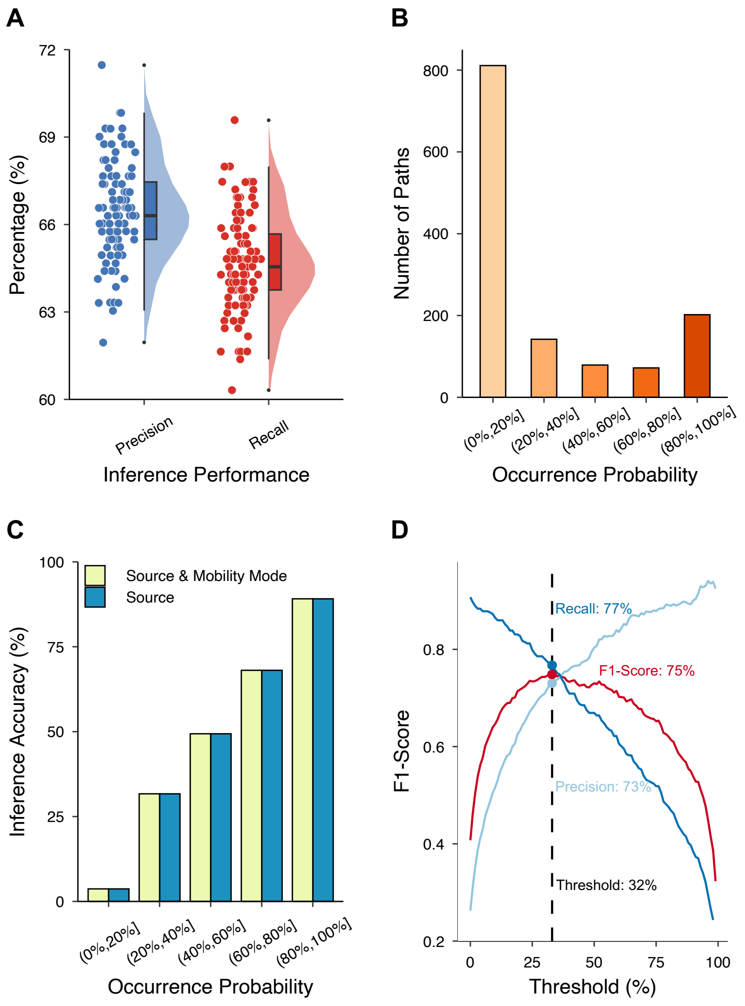


**Fig. S11. Inferring the spatial spread of a simulated outbreak on a randomized mobility network that keeps the mobility volume for each link.** (**A**). Precision and recall for the results from 100 independent realizations of inference. Each dot represents the performance for one realization. Boxes show the medians and interquartile of the distributions and whiskers show 95% CIs. An inferred transmission link is defined as accurate if both the infection source and mobility mode are correct. (**B**). Numbers of transmission links that appeared with different probabilities in 100 realizations of inference. (**C**). The accuracy of inferred transmission links with different occurrence probabilities in the inference. Blue bars show the accuracy for identifying infection sources and yellow bars show the accuracy for additionally finding the correct mobility mode. (**D**). The performance of the ensemble inference using different thresholds of the occurrence probability to define transmission links.

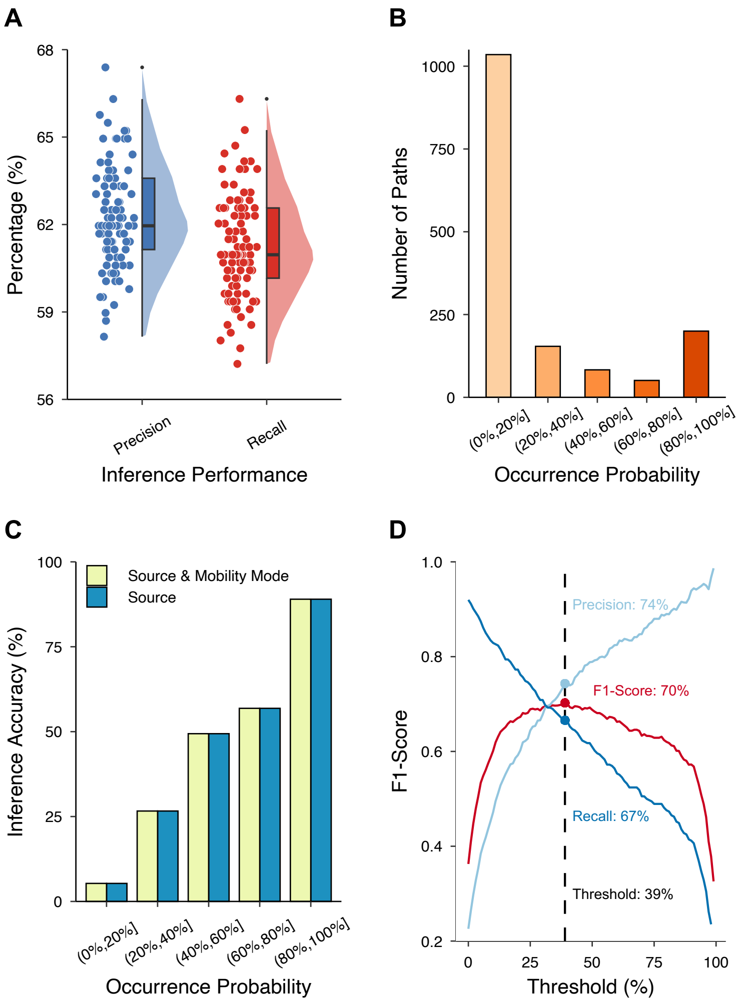


**Fig. S12. Inferring the spatial spread of a simulated outbreak on a randomized mobility network with evenly distributed mobility volume among all commuting or flight links from each MSA .** (**A**). Precision and recall for the results from 100 independent realizations of inference. Each dot represents the performance for one realization. Boxes show the medians and interquartile of the distributions and whiskers show 95% CIs. An inferred transmission link is defined as accurate if both the infection source and mobility mode are correct. (**B**). Numbers of transmission links that appeared with different probabilities in 100 realizations of inference. (**C**). The accuracy of inferred transmission links with different occurrence probabilities in the inference. Blue bars show the accuracy for identifying infection sources and yellow bars show the accuracy for additionally finding the correct mobility mode. (**D**). The performance of the ensemble inference using different thresholds of the occurrence probability to define transmission links.

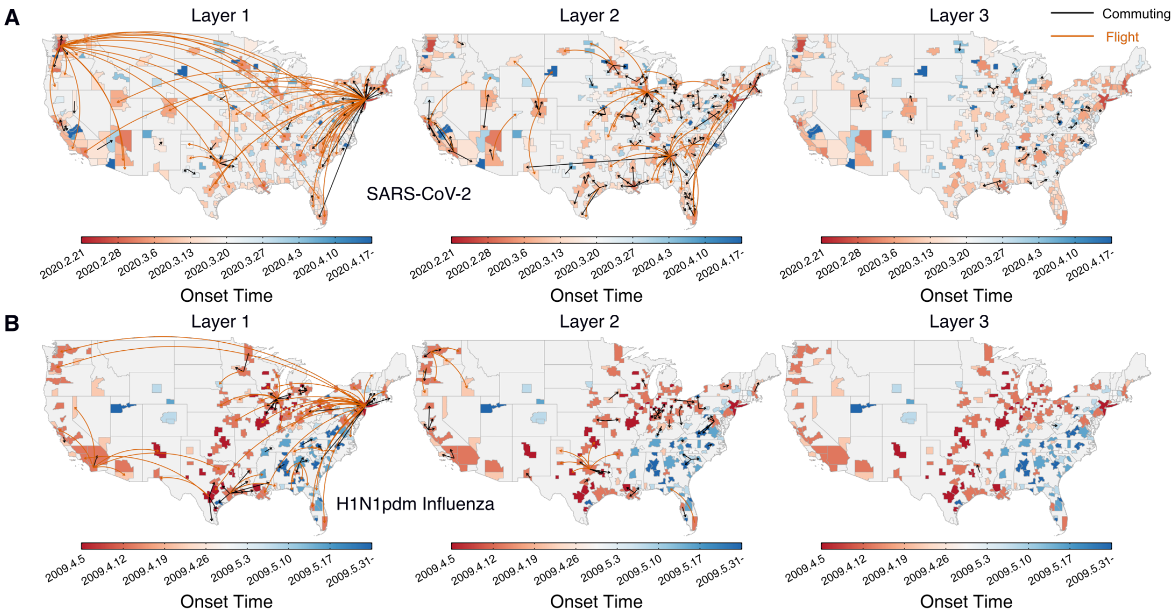


**Fig. S13. The hierarchical structure of reconstructed transmission networks of SARS-CoV-2 and H1N1pdm influenza.** The inferred transmission links of SARS-CoV-2 (**A**) and H1N1pdm influenza (**B**) at different layers from the origin MSAs.

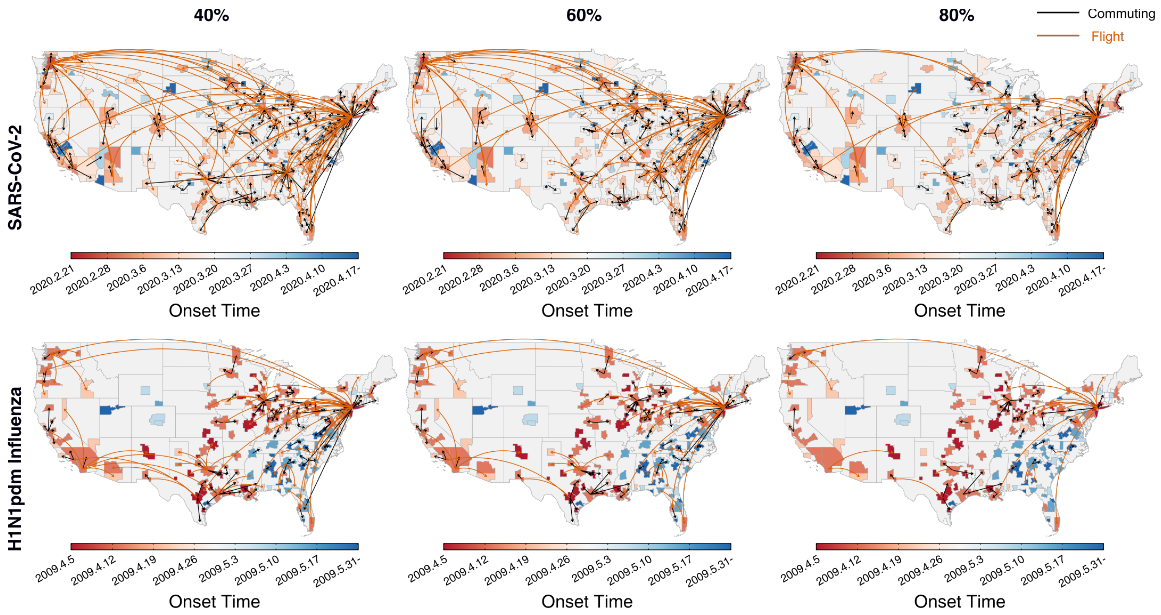


**Fig. S14. The reconstructed transmission networks using different thresholds in the ensemble inference.** Transmission networks for SARS-CoV-2 (upper row) and H1N1pdm influenza (lower row) inferred using a 40%, 60%, or 80% threshold of occurrence probability in the ensemble inference.

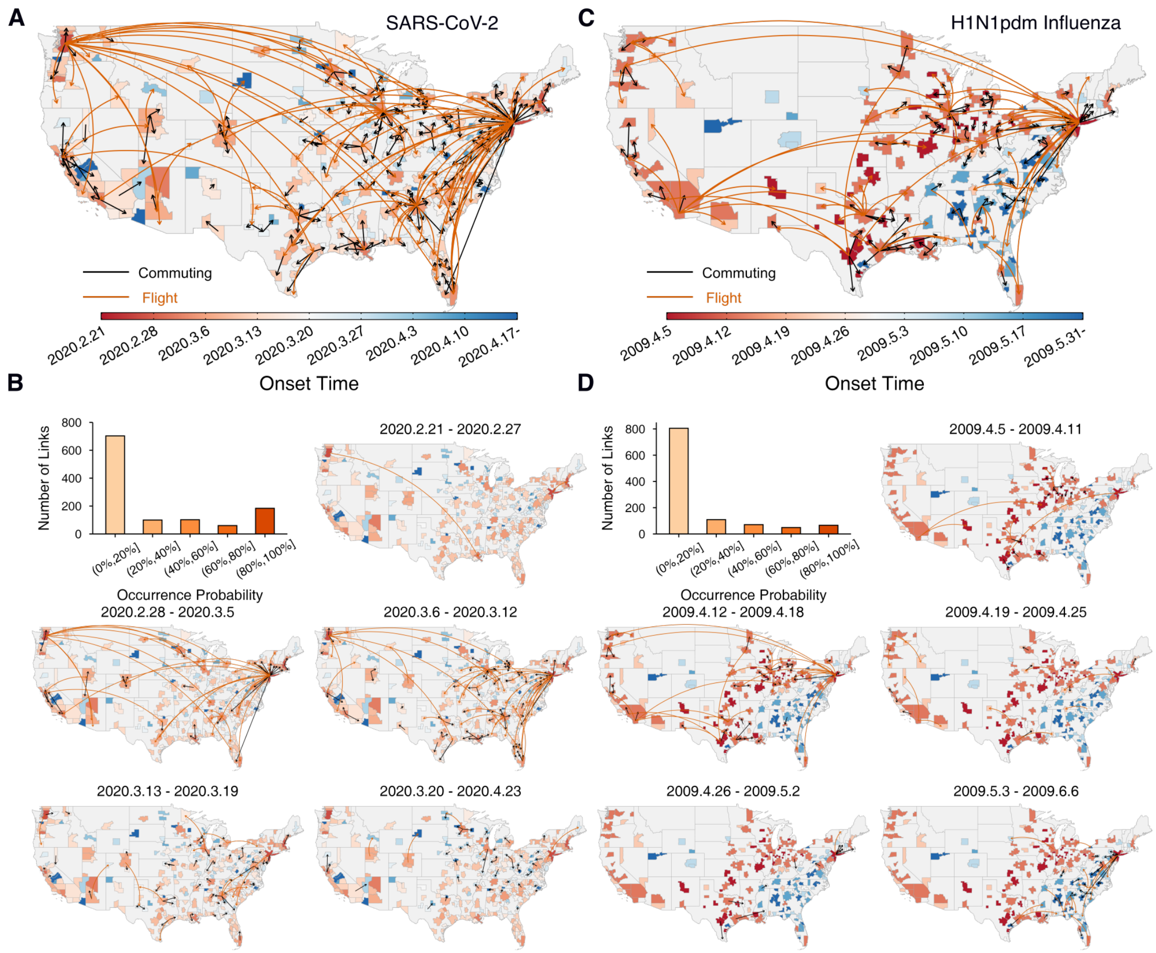


**Fig. S15. Reconstructed transmission networks using alternative set of origin MSAs.** (**A, B**). The reconstructed transmission network of SARS-CoV-2 using three origin MSAs – Seattle–Tacoma–Bellevue, WA MSA, New York-Newark-Jersey City, NY-NJ-PA MSA, and San Francisco-Oakland-Hayward, CA MSA. (**C, D**). The reconstructed transmission network of H1N1pdm influenza using six origin MSAs – New York-Northern New Jersey-Long Island, NY-NJ-PA MSA, San Antonio, TX MSA, San Diego-Carlsbad-San Marcos, CA MSA, Terre Haute, IN MSA, Fort Wayne, IN MSA and Springfield, IL MSA.

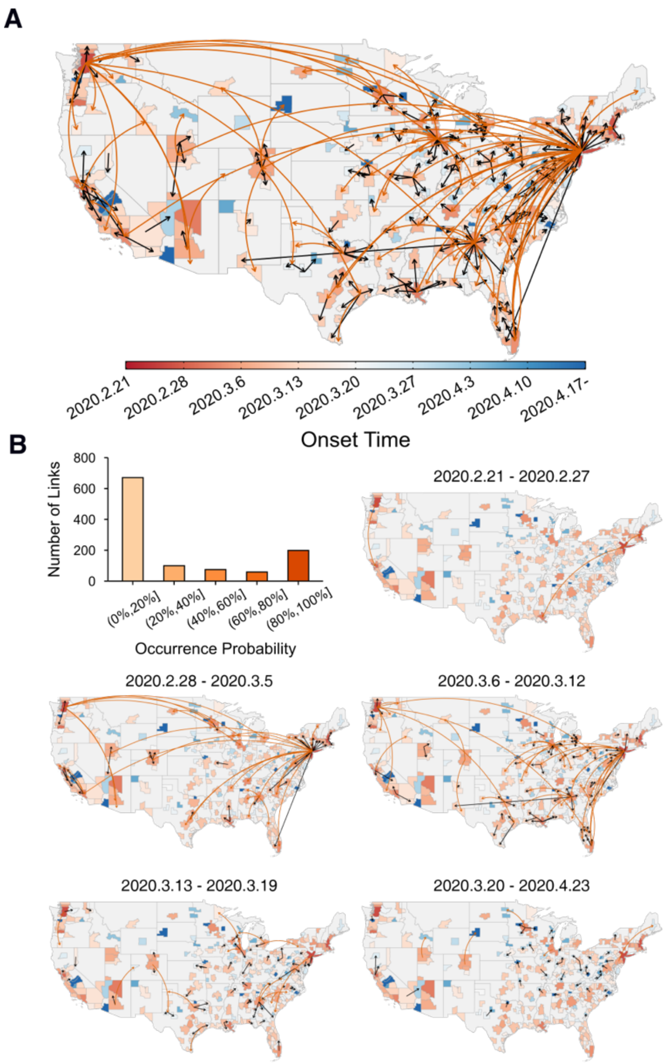


**Fig. S16. Reconstructed transmission networks using a shorter immunity duration.** (**A, B**). The reconstructed transmission network of SARS-CoV-2 using an immunity duration of $L = 0.5$ year.

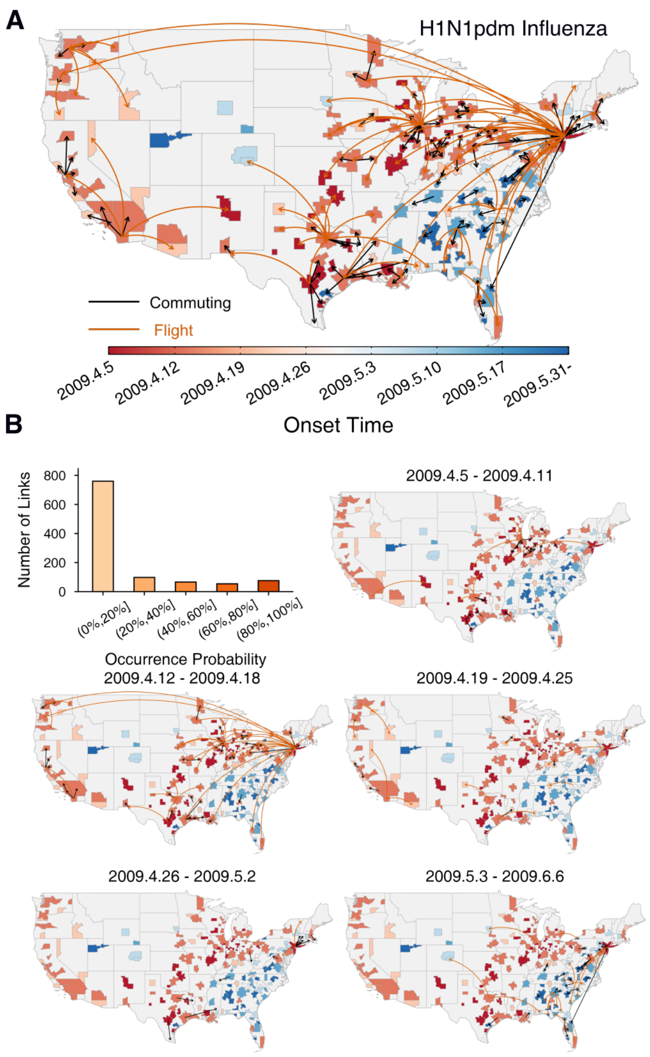


**Fig. S17. Reconstructed transmission networks using different initial infections for H1N1pdm influenza.** (**A, B**). The reconstructed transmission network of H1N1pdm influenza using three origin MSAs – New York-Northern New Jersey-Long Island, NY-NJ-PA MSA, San Antonio, TX MSA, San Diego-Carlsbad-San Marcos, CA MSA. The initial infection number was drawn from $U[100,1000]$.

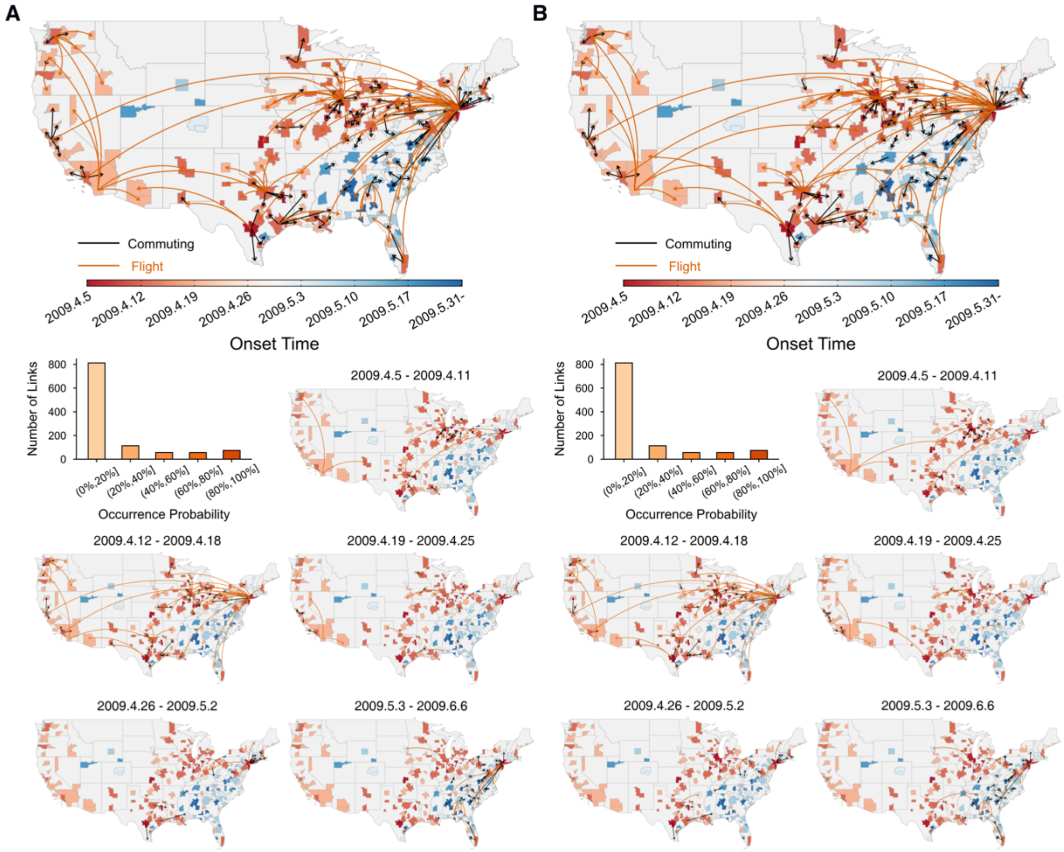


**Fig. S18. Reconstructed transmission networks for H1N1pdm influenza assuming a weekly increase of the detection rate during the first five weeks.** (**A**). Assume a weekly 20% increase of the detection rate during the first five weeks. The ILI+ in each MSA in week $t$ was multiplied by $1/(80\%)^{6-t}$ for $1 \leq t \leq 5$. (B). Assume a weekly 40% increase of the detection rate during the first five weeks. The ILI+ in each MSA in week $t$ was multiplied by $1/(60\%)^{6-t}$ for $1 \leq t \leq 5$.

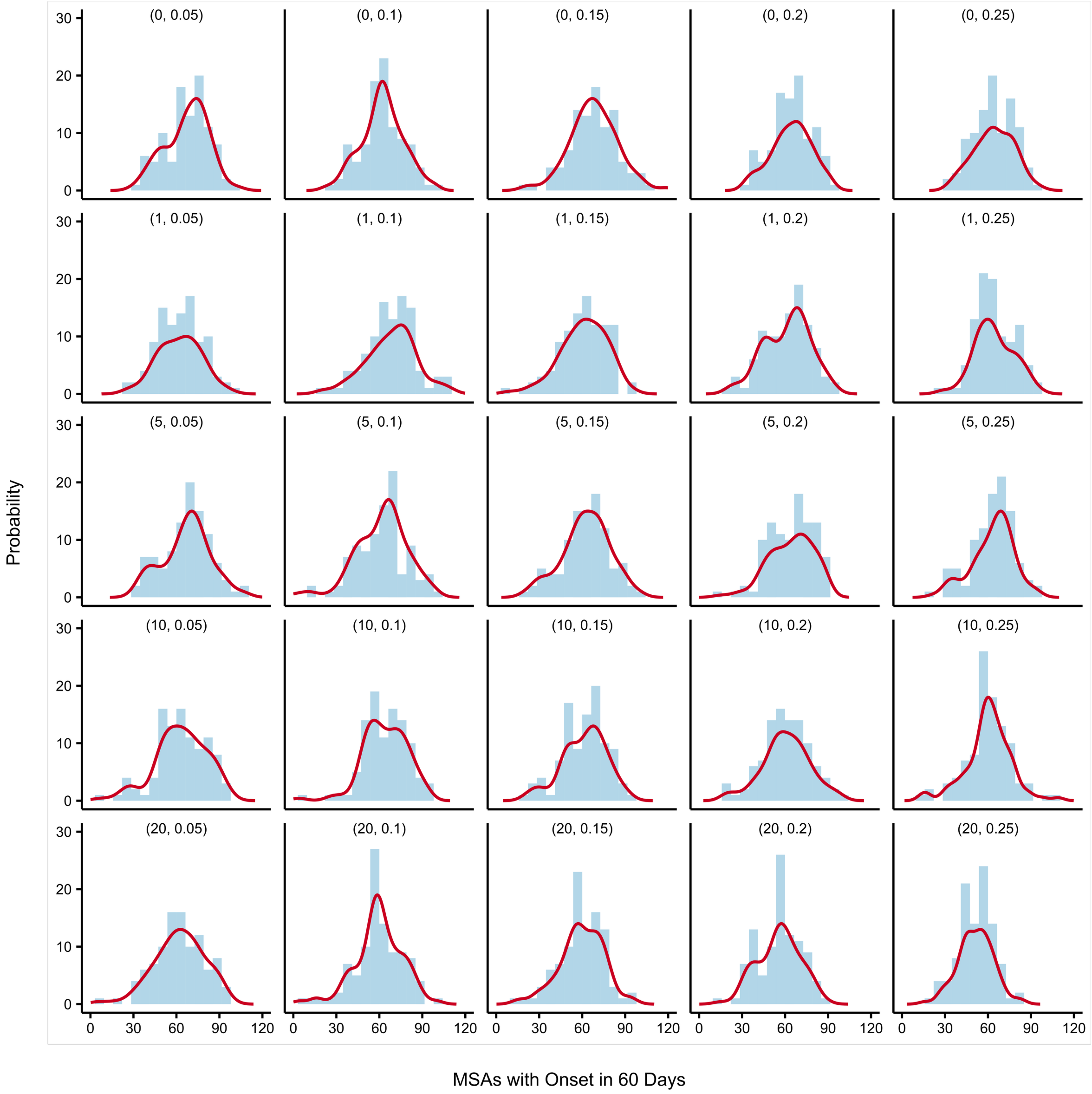


**Fig. S19. The distributions of spatial spread for an outbreak originating from New York under different surveillance and control scenarios.** We simulated an outbreak from New York and examined the number of MSAs with local onsets in the first 60 days under different surveillance and control scenarios. The title $(X, Y)$ shows that the number of transmission hubs with wastewater surveillance is $X$ and the transmission rate reduction is $Y$ if introductions are identified or community infections exceed 5,000 cumulatively. For each scenario, we performed 100 independent simulations. The red lines are the distributions estimated using a Gaussian kernel density estimation.

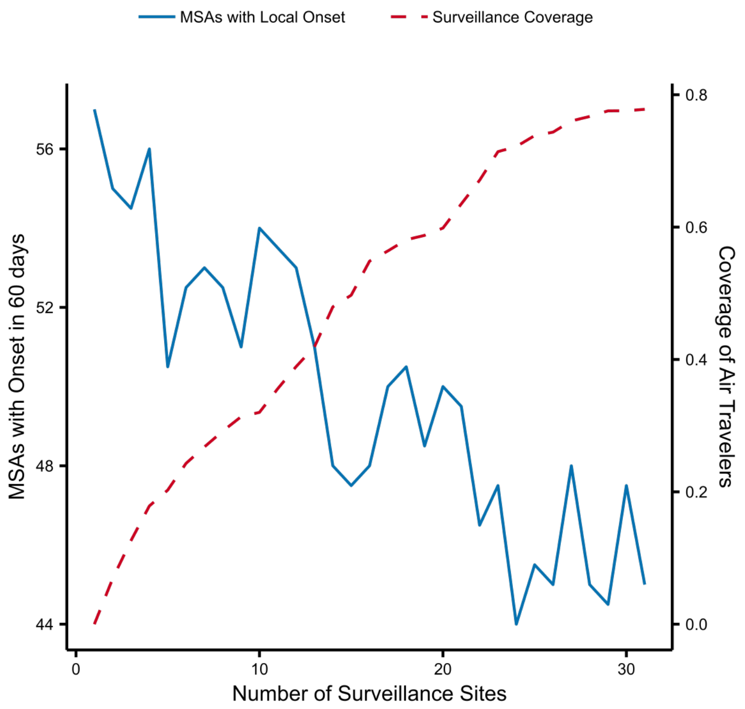


**Fig. S20. Spatial spread for an outbreak originating from New York using a threshold of 1,000 cumulative infections to enact interventions.** The number of MSAs with local onsets in the first 60 days with different coverage of wastewater surveillance is shown. Local transmission rate reduction is enacted when introductions are identified in wastewater surveillance at airports or the cumulative community infections reached 1,000 in an MSA. The transmission rate reduction was fixed at 25% for interventions. The solid line shows the median values obtained from 100 independent simulations. The dash line shows the cumulative coverage of air travelers.

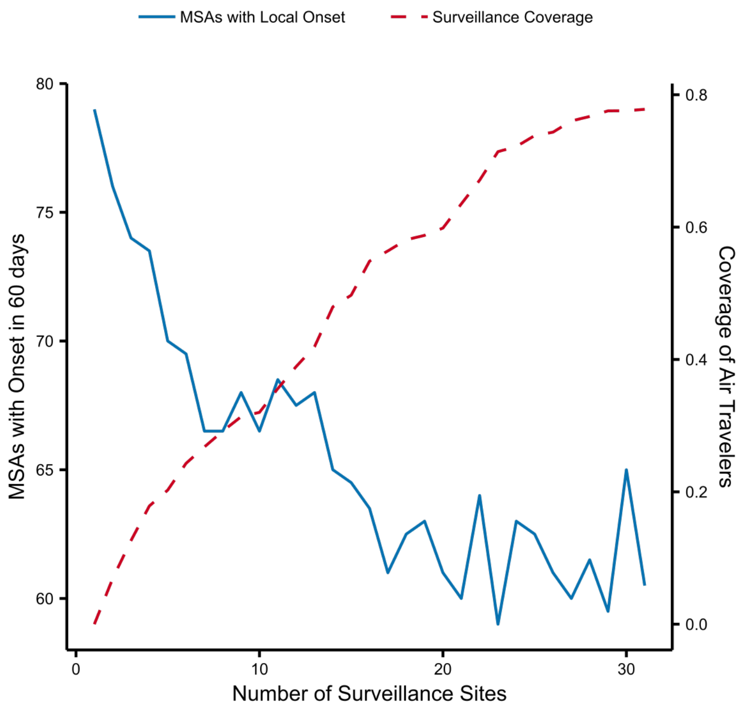


**Fig. S21. Spatial spread for an outbreak originating from Minnesota.** We simulated an outbreak originating from Minnesota using the same configurations in Fig. 1A. The number of MSAs with local onsets in the first 60 days with different coverage of wastewater surveillance is shown. Local transmission rate reduction is enacted when introductions are identified in wastewater surveillance at airports or the cumulative community infections reached 5,000 in an MSA. The transmission rate reduction was fixed at 25% for interventions. The solid line shows the median values obtained from 100 independent simulations. The dash line shows the cumulative coverage of air travelers.

**Table S1. Top 10 MSAs with the largest number of international travelers to other countries in 2009 and 2020.** We computed the annual number of international travelers leaving each MSA to other countries via air travel in 2009 and 2020. Data were from the US Bureau of Transportation Statistics. Note, the names of some MSAs were different in 2009 and 2020.

| MSA | Annul number of international travelers |
|---|---|
| 2009 | |
| New York-Northern New Jersey-Long Island | 8371344 |
| Miami-Fort Lauderdale-Miami Beach | 5933091 |
| Atlanta-Sandy Springs-Marietta | 3920046 |
| Houston-Baytown-Sugar Land | 3153480 |
| Chicago-Naperville-Joliet | 2935706 |
| Dallas-Fort Worth-Arlington | 1983102 |
| Washington-Arlington-Alexandria | 1566460 |
| Philadelphia-Camden-Wilmington | 1554446 |
| Los Angeles-Long Beach-Santa Ana | 1528373 |
| San Francisco-Oakland-Fremont | 1488220 |
| 2020 | |
| Miami-Fort Lauderdale-West Palm Beach | 3329205 |
| New York-Newark-Jersey City | 3178069 |
| Houston-The Woodlands-Sugar Land | 1464867 |
| Dallas-Fort Worth-Arlington | 1422063 |
| Atlanta-Sandy Springs-Roswell | 1405291 |
| Los Angeles-Long Beach-Anaheim | 826987 |
| Chicago-Naperville-Elgin | 767224 |
| San Francisco-Oakland-Hayward | 614577 |
| Charlotte-Concord-Gastonia | 506958 |
| Minneapolis-St. Paul-Bloomington | 389245 |

**Table S2. The selected outbreak origins in simulations.** For each simulation, we selected one outbreak origin in the US. The left column shows the origins within the US, informed by the number of detected HPAI spillovers. The right column shows the locations with intense international travels as potential points of introduction of novel pathogens into the US.

| Originating within US | Introduced into US |
|---|---|
| Phoenix-Mesa-Scottsdale, AZ MSA | San Francisco-Oakland-Fremont, CA MSA |
| Fort Collins-Loveland, CO MSA | Los Angeles-Long Beach-Santa Ana, CA MSA |
| Wichita, KS MSA | San Diego-Carlsbad-San Marcos, CA MSA |
| Baltimore-Towson, MD MSA | Seattle-Tacoma-Bellevue, WA MSA |
| Lansing-East Lansing, MI MSA | Portland-Vancouver-Beaverton, OR-WA MSA |
| Minneapolis-St. Paul-Bloomington, MN-WI MSA | Boston-Cambridge-Quincy, MA-NH MSA |
| St. Louis, MO-IL MSA | New York-Northern New Jersey-Long Island, NY-NJ-PA MSA |
| Albuquerque, NM MSA | Philadelphia-Camden-Wilmington, PA-NJ-DE-MD MSA |
| Lancaster, PA MSA | Miami-Fort Lauderdale-Miami Beach, FL MSA |
| Memphis, TN-MS-AR MSA | Orlando, FL MSA |
